# Homologous recombination deficiency in primary ER-positive and HER2-negative breast cancer

**DOI:** 10.1101/2025.06.04.25328880

**Authors:** Helen R. Davies, Daniella Black, Anders Kvist, Kristín Sigurjónsdóttir, Ana Bosch Campos, Ramsay Bowden, Yasin Memari, Ziqian Chen, Giuseppe Rinaldi, Frida Rosengren, Deborah F. Nacer, Srinivas Veerla, Lennart Hohmann, Nicklas Nordborg, Jari Häkkinen, Johan Vallon-Christersson, Åke Borg, Serena Nik-Zainal, Johan Staaf

## Abstract

Homologous recombination deficiency (HRD) originating from inactivation of genes like *BRCA1*/*BRCA2* is a targetable abnormality common in triple-negative breast cancer (TNBC). In estrogen-receptor (ER)-positive HER2-negative (ERpHER2n) breast cancer (BC), HRD prevalence and clinical impact are unclear. We analyzed 502 ERpHER2n tumors from patients recruited via the population-representative Swedish SCAN-B study, by whole genome sequencing (WGS) defining mutational signatures-based HRD, as well as matched transcriptional, DNA methylation, clinicopathological, treatment and outcome data. HRD is much less frequent in ERpHER2n BC (8.4%) compared to TNBC (58.6%), though induced by similar genetic/epigenetic mechanisms acting on mainly *BRCA1*/*BRCA2*/*RAD51C*/*PALB2*. Our modelled estimate of HRD in Western European BC is ∼10-13%. HRD tumors were observed across all gene expression subtypes and did not exhibit a unique, defining transcriptional or DNA methylation profile. Though numbers are limiting, we present early evidence that HRD stratification by WGS could impact therapeutic strategies, as HRD BCs trended to poorer outcomes, especially when not treated with chemotherapy.

## INTRODUCTION

Breast cancers (BC) that are estrogen receptor (ER) positive and human epidermal growth factor receptor 2 (HER2) negative (ERpHER2n) constitute the largest clinical subgroup of primary BC, representing ∼70% of all BC cases [1]. The ERpHER2n subgroup is a heterogeneous mix of tumors with varying clinical aggressiveness, treated typically along two main lines: adjuvant endocrine therapy only (Endo) or combined neo/adjuvant systemic chemotherapy and endocrine therapy (ChemoEndo). Current treatment guidelines now suggest complementing ChemoEndo treatment with CDK4/6 inhibitors or potentially PARP-inhibitors depending on *BRCA1/2* germline status for high-risk patients (see e.g. [2]). Despite continuous improvements in therapeutic strategies, many patients with primary ERpHER2n BC relapse, often many years following the original diagnosis. This represents a substantial clinical challenge requiring improved methods of risk assessment and identification of new targets for selective therapeutics. For decades, immunohistochemistry and diverse genomic and transcriptomic methods have been explored in ERpHER2n BC, resulting in an array of reported prognostic and/or predictive biomarkers [3, 4]. Only a handful have reached clinical implementation and are proven to aid therapeutic decisions in restricted tumor subsets [2, 5].

In parallel to the extensive transcriptional characterization of BC, large scale whole genome sequencing (WGS) studies have delineated the genomic landscape of BC, pinpointing not only specific oncogenic drivers of disease but also the broader mutational landscape inferred by mutational processes (e.g. [6]). One prominent mutational process is homologous recombination (HR) deficiency (HRD), typically caused by disruption of genes involved in the HR pathway, e.g., *BRCA1*, *BRCA2, PALB2* and *RAD51C*, by various mechanisms including germline and/or somatic mutation and promoter hypermethylation. Tumor HRD classification is based on genomic analyses [7], historically using copy number aberrations or targeted panels, and more recently through whole genome sequencing (WGS) approaches. HRD confers specific patterns of somatic mutations and structural variation (referred to as mutational signatures) [6] forming the foundation of WGS-based predictors such as HRDetect [8]. Applications of HRD predictors have further unveiled that a substantial proportion of BC displays somatic signatures of HRD, with reports of close to 60% in triple negative breast cancers (TNBCs), some without known pathogenic germline or somatic variants and/or promoter hypermethylation of HR genes [8, 9]. Critically, tumors with HRD signatures have been shown to respond favorably to compounds that increase the demand on compensatory repair pathways such as DNA-damaging agents (e.g., platinum) and PARP-inhibitors, irrespective of HR gene status (e.g., [10–14]), however with strongest clinical trial data support mainly in ovarian cancer [15, 16].

In primary BC, current guidelines recommend adjuvant PARP-inhibitor therapy for patients with pathogenic germline variants (PGVs) in *BRCA1/BRCA2*, but not yet for tumors with an HRD phenotype per se [2]. In part, this may be due to conflicting results about the association of HRD with treatment response caused by the variation in how HRD is defined and measured (see [17, 18]). HRD classification may not necessarily correlate to active HRD in a tumor, as reports have shown that prolonged treatment with platinum-based or PARP-inhibitor therapies can result in the selection of resistance-causing reversion mutations or gene reactivation by promoter demethylation restoring HR [19, 20]. However, functional HRD assays, like the RAD51 assay, also have multiple practical limitations for clinical use (see [7]), and the degree of concordance of HRD status by genetic classifiers and functional HRD assays in primary treatment-naïve patients, while promising, is not fully established [11].

Unlike in TNBC, the landscape of HRD in the most common BC subtype of ERpHER2n remains unclear. In the earliest days of next-generation sequencing, tumors that were sequenced were not representative of a more general patient population [8, 21–23]. Moreover, systematic clinical data collection was lacking and thus the relationship between HRD status, treatment, and patient outcome, especially on standard-of-care (SOC) therapy, has not been robustly interrogated. Finally, varying frequencies of HRD-positivity in BC have been reported [8, 22, 24], dependent both on differences in HRD-calling approach and cohort compositions. In this study, we investigate the largest cohort of WGS ERpHER2n BC (n=502) within a SOC therapy setting involving adjuvant chemotherapy and endocrine therapy by taking advantage of the prospective, population-representative, Sweden Cancerome Analysis Network – Breast (SCAN-B) study [25] providing long term clinical follow-up. In addition to WGS, RNA-sequencing, global DNA methylation, and complete clinical follow-up data are also included. Here, we investigate the frequency, causes, molecular associations, and prognostic implications of HRD in primary ERpHER2n BC collected in a routine diagnostic setting.

## RESULTS

### HRD frequencies and relationship with transcriptional subtypes

We first evaluated the population representativeness of our WGS-analyzed primary SCAN-B ERpHER2n BC cohort (with no patients receiving treatment prior to tissue sampling). Indeed, ChemoEndo cases in our study were comparable to ChemoEndo patients in the complete catchment region for inclusion years, while the Endo group in our study was skewed towards a more aggressive phenotype, as shown by e.g., higher proportions of grade 3 tumors and more proliferative tumors according to the Ki67 marker, consistent with the study’s patient selection criteria and layout (see **Supplementary Figure S1A-D**, **Supplementary Methods**).

Of the 502 patients, 42 (8.4%) were classified as HRD (HRDetect score ≥0.9, otherwise HR-proficient), with an HRD frequency of 7.2% in the Endo and 9.1% in the ChemoEndo treatment group (**Figure 1A**, **Table 1, Supplementary Table S1**). Of the 42 patients, three (7.1%) had a previously known germline pathogenic variant in *BRCA1/BRCA2*, three (7.1%) had somatic homozygous *BRCA1*/*BRCA2* deletions, and 10 (23.8%) had only somatic pathogenic variants, indels, or structural rearrangements affecting *BRCA1*, *BRCA2*, or *PALB2* detected by WGS. An HRDetect-intermediate classification (score ≥0.1 and <0.9) was observed in 8.6% of patients. Patients with HRD tumors showed no significant difference in age at diagnosis, tumor size, HER2-low status, and lymph node status compared to HR-proficient cases for the total cohort, the ChemoEndo subset, or the Endo subset (p>0.05, Wilcoxon’s test or Chi-square test). HRD tumors showed higher Ki67-index, lower levels of ER and PR-stained cells, and higher tumor grade (non-significant trend in Endo subset) compared to other tumors in the total cohort, ChemoEndo and/or Endo subsets (**Supplementary Figure S2**). Two tumors (0.4%) showed signatures of mismatch repair deficiency (MMRd). Both were scored as HR-proficient by HRDetect, however, one (S002375), with a somatic biallelic *BRAC2* mutation, appears to have a mixed HRD/MMRd phenotype. Finally, six of the 502 patients were male, only one with a tumor classified as HRD (16.7%).

**Figure 1.**
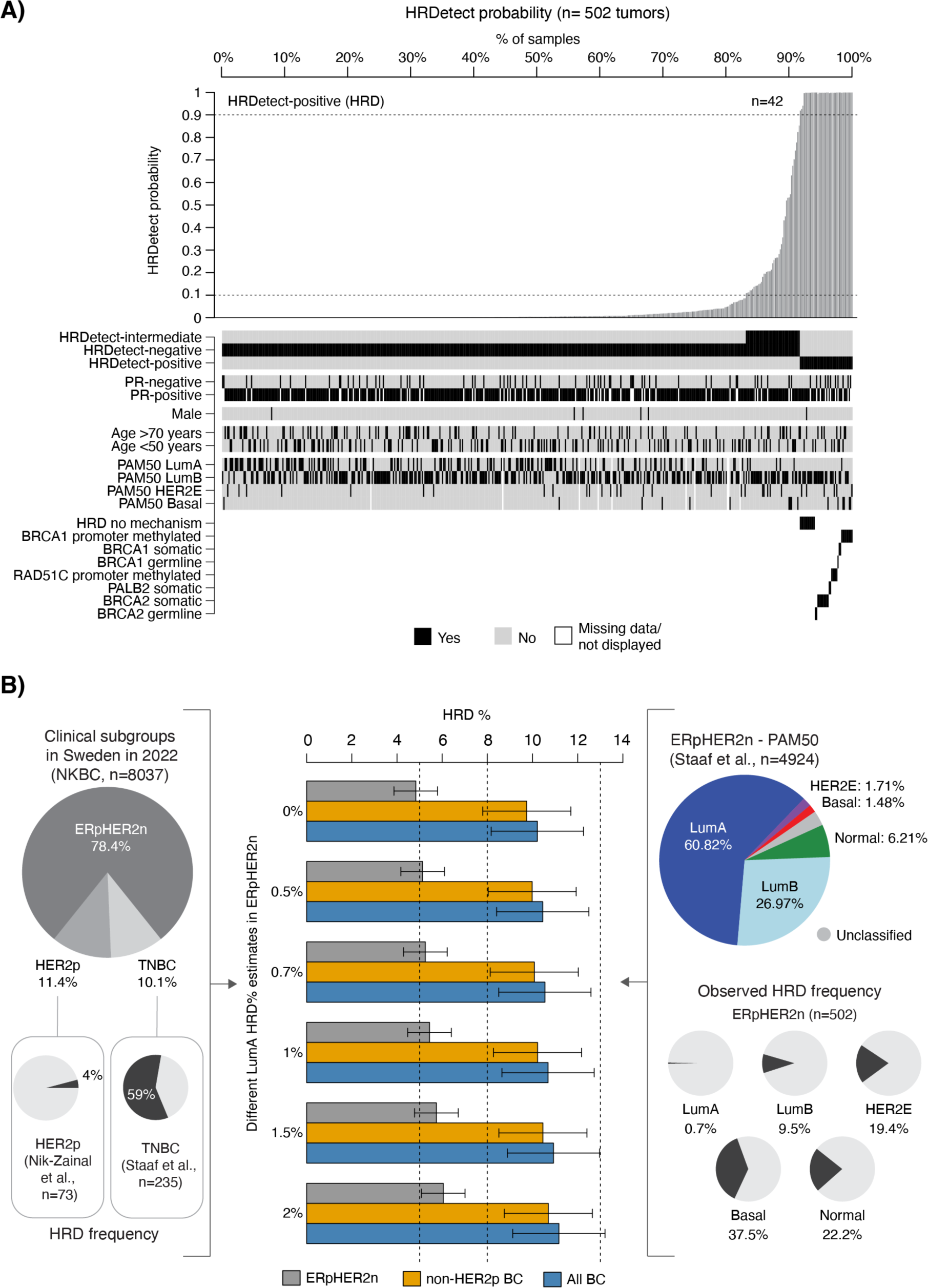
HRD in ERpHER2n. (**A**) HRDetect probability for 502 WGS analyzed ERpHER2n SCAN-B tumors and their characteristics. HRD is defined as an HRDetect probability ≥0.9. (**B**) Estimation of HRD frequency in breast cancer for ERpHER2n tumors, non-HER2-positive tumors (non-HER2p) and all breast cancer (including ERpHER2n, HER2-positive, and TNBC tumors). Estimations are based on the clinical subgroup proportions reported to the national Swedish breast cancer registry (NKBC) in 2022 (left pie chart), the reported PAM50 subtype proportions in ERpHER2n tumors from [28] (right pie chart), HRD frequency in population-based TNBC [9], HRD frequency in HER2-positive disease [6], and the estimated HRD frequency in PAM50 Basal, HER2E, LumB, and Normal subtypes in this study. Final estimates are presented as a range based on calculations using different assumed HRD frequencies in ERpHER2n LumA tumors. Error bars represent the span of estimated HRD frequency based on assumption of 20% error in the individual HRD point estimates.

**Table 1.** Clinicopathological characteristics of the ERpHER2n WGS cohort and total HRD frequency.

|  | Total cohort | ChemoEndo<sup>1</sup> | Endo |
| --- | --- | --- | --- |
| Number of patients | 502 (100%) | 339 (67.5%) | 138 (27.5%) |
| Female / male % | 98.8/1.2 % | 99.7/0.3% | 97.1/2.9 % |
| Median age (years) | 65 | 60 | 75 |
| Median tumor size and range (mm) | 20 (2-115) | 20 (2-115) | 18 (8-67) |
| Nottingham Grade (NHG) |  |  |  |
| NHG 1 | 8.1% | 6.4% | 10.1% |
| NHG 2 | 40.0% | 35.8% | 49.3% |
| NHG 3 | 51.8% | 57.9% | 40.6% |
| Nodal status |  |  |  |
| Node-negative (N0) | 47.3% | 38.4% | 62.3% |
| Node-positive (N+) | 52.7% | 61.6% | 37.7% |
| HER2-low status<sup>2</sup> | 84.8% | 84.6% | 85.7% |
| PR-status (positive/negative) | 84.8/15.2% | 86.1/13.9% | 81.9/18.1% |
| Adjuvant therapy |  |  |  |
| ChemoEndo<sup>1</sup> | 71.1% | 100% | 0% |
| Endo | 28.9% | 0% | 100% |
| Complete overall survival data | 100% | 100% | 100% |
| Complete DRFI survival data<sup>3</sup> | 99.6% | 98.5% | 98.6% |
| HRD frequency | 8.4% | 9.1% | 7.2% |
| PAM50 subtypes |  |  |  |
| Basal | 3.3% | 2.1% | 4.3% |
| HER2-enriched (HER2E) | 6.3% | 4.0% | 11.6% |
| Luminal A (LumA) | 30.9% | 37.1% | 18.1% |
| Luminal B (LumB) | 57.7% | 54.1% | 65.9% |
| Normal | 1.8% | 2.7% | 0% |

Note: Cases with missing values for a variable are excluded from percentage calculations if not otherwise stated.
<sup>1</sup> Only ChemoEndo cases with clinical review data are included.
<sup>2</sup> HER2-low classification was only possible in reviewed cases (including some, but not all, Endo cases). Cases with missing values are omitted.
<sup>3</sup> By clinical review or cancer registry data depending on cohort.

We next assessed the relationship between HRD in ERpHER2n tumors and PAM50 subtypes, a common BC stratification strategy based on gene expression. HRD frequencies were 37.5% in the PAM50 Basal subtype (6/16), 19.4% in HER2-enriched (HER2E) (6/31), 0.7% in Luminal A (LumA) (1/152), 9.5% in Luminal B (LumB) (27/284) tumors and 22.2% in Normal (2/9) tumors. Thus, HRD can occur across all PAM50 subtypes, albeit with different frequencies.

### Population frequency of HRD in ERpHER2n BC

For each PAM50 subtype, we further contrasted patient age, tumor size, lymph node status, tumor grade, and gene expression rank scores of five biological metagenes (representing immune response, steroid response, basal expression, and proliferation) [26, 27] between our WGS cohort versus an independent set of 4,427 non-WGS ERpHER2n tumors of unknown HRD status [28] (**Supplementary Methods**). These analyses demonstrated that the PAM50 Basal, HER2E, and LumB subgroups in the WGS cohort showed similar characteristics to non-WGS profiled cases and may be viewed as population-representative, whereas PAM50 LumA and Normal tumors in this WGS cohort cannot be considered population-representative (**Supplementary Figure S3**).

Based on these results, we estimate the prevalence of HRD in ERpHER2n BC and BC as a whole by combining our estimates with HRD estimates in TNBC and HER2-positive tumors from previous publications [6, 9]. Specifically, we merged our HRD estimations in the PAM50 Basal, HER2E, LumB, and Normal subtypes with additional population-representative BC datasets reporting PAM50 subtype proportions in ERpHER2n tumors (n=4,924 tumors) [28], HRD frequency in TNBC (59%) [9], HRD frequency in HER2-positive tumors [6] (4.1%, 4/73 WGS and HRDetect analyzed cases), and data from the Swedish National Breast Cancer Quality Registry (NKBC) of the population proportions of TNBC, HER2-positive, and ERpHER2n disease in Sweden 2022 (**Figure 1B**). Combining HRD and PAM50 subtype proportions for ERpHER2n tumors we infer a HRD frequency of 4.8% in ERpHER2n BC (excluding the contribution from LumA or unclassified tumors). Next, combining the ERpHER2n HRD estimate with NKBC clinical subtype proportions and our HRD estimates in TNBC and HER2-positive tumors allows us to model an overall HRD frequency that varies depending on assumptions of HRD proportions in ERpHER2n LumA tumors (excluding unknown contributions from the small set of PAM50 unclassified ERpHER2n tumors in [28]). For instance, with the observed HRD frequency of 0.7% in ERpHER2n LumA tumors, the final overall HRD frequency would be 10.1%, or 1 in 10 in HER2-negative BC patients, or 10.5% in all breast cancer (TNBC, HER2-positive, and ERpHER2n combined).

### Agreement in HRD classifications by alternative methods in ERpHER2n BC

HRD status can be predicted in multiple ways using DNA-based methods [7, 17, 18]. To assess the agreement of different DNA-based methods we compared HRD classification by HRDetect versus HRD status called by CHORD (Classifier of HOmologous Recombination Deficiency) [24], scarHRD (representing an HRD score involving telomeric allelic imbalances, loss of heterozygosity profiles, and large-scale state transitions) [29], and a copy number signature approach (based on the CN17 signature proposed to be associated with HRD by Steele et al. [30]). For scarHRD, we found a concordance of 0.94 in HRD status compared to HRDetect; however, only about 60% of scarHRD HRD tumors were HRD by HRDetect resulting in a lower positive predictive value (PPV=0.58) (**Figure 2A**). Similarly, while CN17 proportions were significantly associated with HRDetect status (**Figure 2B**), thresholded classification of CN17 proportions showed that about 55-60% of CN17 HRD cases were HRD by HRDetect (**Figure 2C**). For the sequencing-based CHORD method, 466 of the 502 tumors could be classified (92.8%). Agreement between CHORD and HRDetect for these 466 tumors was 0.99, while the PPV was 0.89 (**Figure 2D**). Venn diagrams for HRD and HR-proficient tumors revealed that 27 of the 42 (64.2%) HRDetect HRD tumors were classified as HRD by all four methods, while 82.4% (379/460) of HRDetect HR-proficient tumors were classified as HR-proficient by all methods (**Figure 2E**).

**Figure 2.**
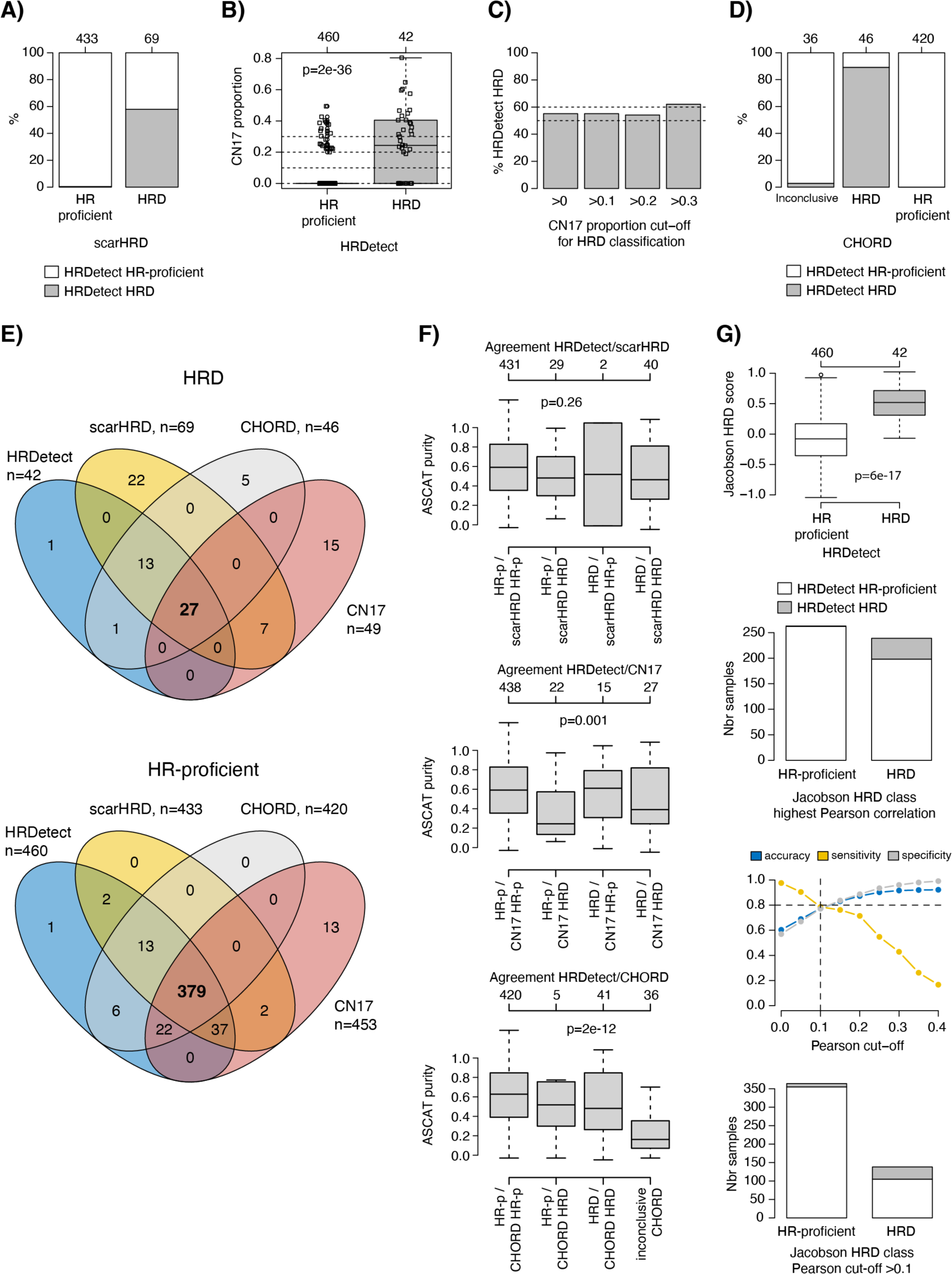
Comparison of DNA-based HRD classification methods in ERpHER2n. (**A**) HRDetect status for tumors classified as scarHRD HR-proficient or HRD. (**B**) CN17 signature proportions versus HRDetect status. Two-sided p-values calculated using Wilcoxon’s test. (**C**) Proportion of HRDetect HRD tumors in groups of CN17 HRD tumors classified using different cut-offs for the CN17 signature proportion. (**D**) HRDetect status for tumors classified as CHORD HR-proficient, HRD, or undetermined. (**E**) Venn diagrams of classification overlap between HRD methods. For CN17 a cut-off of >0 was used to call HRD. For CHORD, 36 tumors could not be classified and are not included. (**F**) ASCAT WGS estimated tumor purities versus concordant and discordant HRD status between HRDetect and the other methods. HR-p: HR-proficient. For CN17 a cut-off of >0 was used to call HRD. Two-sided p-values calculated using Kruskal-Wallis test. (**G**) Results of a gene expression HRD classifier versus HRDetect in the 502 tumors. Panels show from top to bottom: i) distribution of an expression-based HRD score versus HRDetect status with p-value calculated using Wilcoxon’s test, ii) composition of HRDetect HRD status in HRD groups defined by gene expression using highest centroid correlation only, iii) accuracy, sensitivity, and specificity for gene expression vs HRDetect classification using different Pearson correlation cut-offs applied to the HRD centroid, and iv) composition of HRDetect HRD status in HRD groups defined by gene expression using an optimized Pearson cut-off of 0.1. Boxplot elements correspond to: i) center line = median, ii) box limits = upper and lower quartiles, iii) whiskers = 1.5x interquartile range. In boxplots, top axes indicate group sizes.

To assess whether discordance between HRDetect and the other DNA-based methods was due to tumor cellularity we compared ASCAT tumor purity estimates versus combined classifications. This analysis showed that discordance for scarHRD did not seem associated with tumor purity, while tumors that could not be classified by CHORD as well as false positive CN17 tumors (HRDetect HR-proficient but CN17 HRD) appeared to have lower tumor cell content (**Figure 2F**).

Finally, we also compared HRDetect status to HRD status based on a gene expression-based nearest centroid HRD classifier reported by Jacobson et al. [31] for the 502 tumors (**Figure 2G**). While the proposed gene expression HRD score was significantly different between HRDetect HRD and HR-proficient tumors, classification using highest centroid correlation resulted in a substantial number of HRDetect HR-proficient tumors being called as HRD. Instead, analysis of a range of correlation cut-off values suggested that 0.1 was the optimal Pearson correlation cut-off to the HRD centroid (**Figure 2G**). This cut-off resulted in an accuracy, sensitivity, and specificity of HRD of approximately 80% compared to HRDetect, in line with the original study results [31]. Still, this optimized cut-off resulted in a substantial number of HRDetect HR-proficient tumors being called as HRD (**Figure 2G**).

### Transcriptional features of HR-deficient ERpHER2n BC are diverse

HRD tumors have reported selective therapeutic vulnerabilities, raising the question of whether transcriptional profiling could identify HRD for precision medicine purposes. We thus investigated whether ERpHER2n HRD tumors showed consistency in transcriptional profiles across PAM50 subtypes, limiting our analyses to the Basal, HER2E, and LumB PAM50 subtypes where we were powered and more population representative. First, we performed unsupervised gene expression analysis using principal component analysis (PCA) contrasting cases with and without HRD. Variance within tested groups using 5000 of the most variable genes, whether across the whole cohort or separated by PAM50 subtypes, could not be explained by HRD status (**Figure 3A**). Thus, HRD status is not singularly a major determinant of transcriptional variation in ERpHER2n disease.

**Figure 3.**
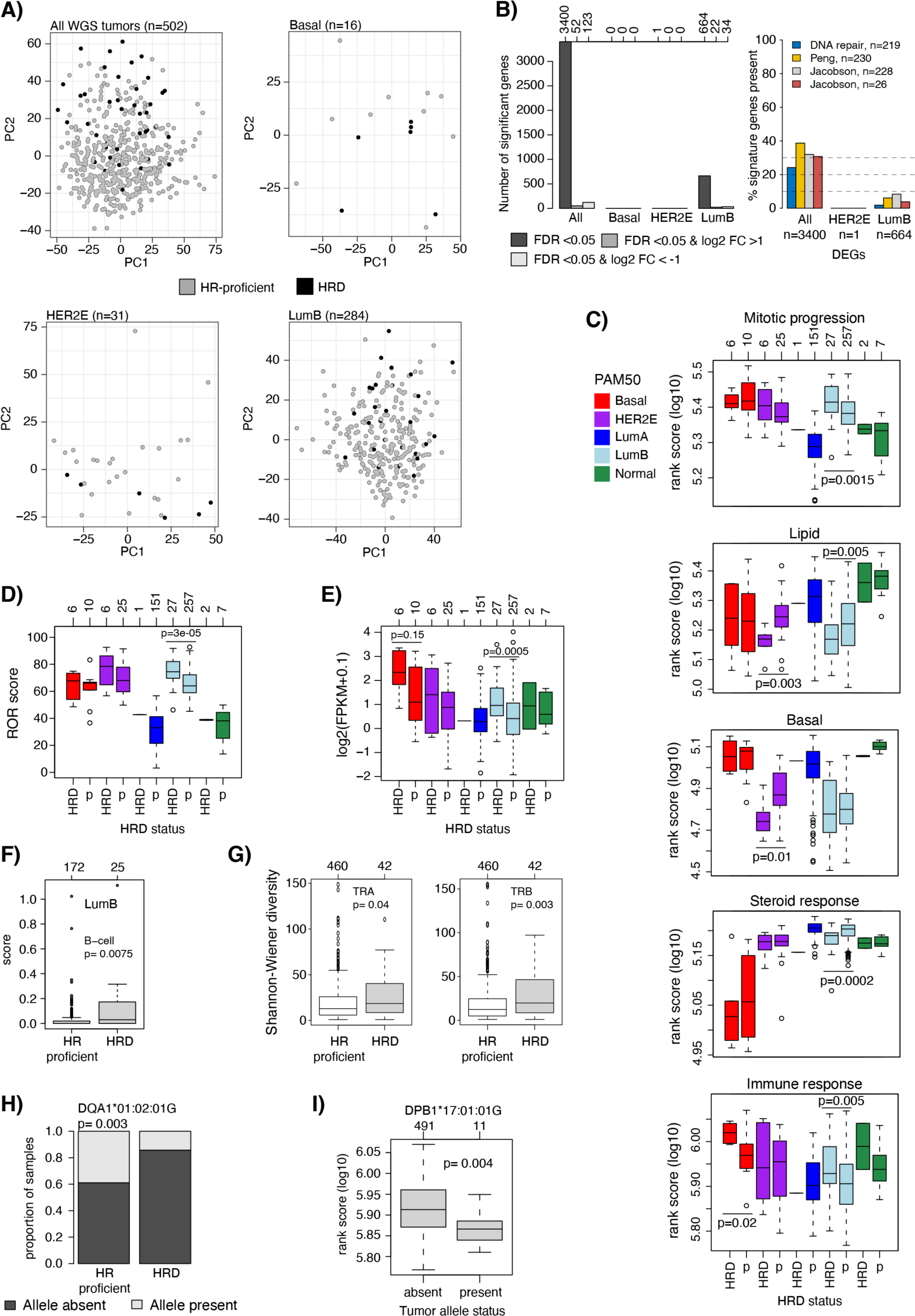
Transcriptional patterns in ERpHER2n BC with respect to HRD status. (**A**) Principal component analysis in all ERpHER2n tumors and in each PAM50 subtype using the 5000 most variant genes based on FPKM data for respective group. HRD tumors are marked by black points. Principal component 1 and 2 (PC1 and PC2, respectively) are shown. The LumA group is excluded due to only one HRD tumor. (**B**) Left: summary of the number of differentially expressed genes per PAM50 subtype based on different FDR-adjusted p-values and log2 fold-changes (FC). Top axis indicates exact number of genes for each bar. Right: proportion of gene overlap between four gene signatures and the differentially expressed gene sets for all tumors, HER2E, and LumB. (**C**) Gene expression rank scores (log10-transformed) for five biological metagenes defined by Fredlund et al. [27] stratified by PAM50 subtype and HRD status (p=HR-proficient). Only significant associations are shown as P-values, calculated using Wilcoxon’s test. Top axis indicates group sizes. (**D**) ROR-score as calculated by Staaf et al. [28]. Two-sided p-values calculated using Wilcoxon’s test. p=HR-proficient. (**E**) mRNA expression of PD-L1 (*CD274*) versus HRD status in PAM50 subtypes. Only significant or borderline non-significant associations are shown as two-sided p-values, calculated using Wilcoxon’s test. p=HR-proficient. (**F**) CIBERSORTx estimates for B-cells in LumB tumors stratified by HRD status. Two-sided p-values calculated using Wilcoxon’s test. Not all tumors have values based on CIBERSORTx p-value filtering. (**G**) Shannon-Wiener diversity scores computed from bulk tumor RNA-sequencing data in all tumors for the T-cell receptor genes *TRA* and *TRB* stratified by HRD status. Two-sided p-values calculated using Wilcoxon’s test. (**H**) Proportion of HR-proficient and HRD tumors with the DQA1*01:02:01G allele present or absent. Two-sided p-value calculated using the Chi-square test. (**I**) Log10 immune rank scores versus present/absent allele status for the DPB1*17:01:01G allele. Two-sided p-value calculated using Wilcoxon’s test. Boxplot elements correspond to: i) center line = median, ii) box limits = upper and lower quartiles, iii) whiskers = 1.5x interquartile range. In boxplots, top axes indicate group sizes.

Second, performing a supervised differentially expressed gene (DEG) analysis contrasting HRD status in the whole cohort, we found that 3,400 genes differed between HRD and HR-proficient tumors. Gene Set Enrichment Analysis (GSEA) using hallmark signatures from MSigDB [32] of this gene set identified 10 significant hallmarks related to proliferation (checkpoint and mitotic spindle), immune response (interferon response, allograft rejection), estrogen signaling, MTORC1 signaling, MYC and E2F targets (adjusted two-sided p<0.05) (**Supplementary Table S2**). Notably, these hallmark enrichments are highly consistent with findings by Ballot et al. [22]. However, when a supervised approach was applied within the Basal, HER2E, and LumB PAM50 subtypes, no DEGs were detected in Basal and only one gene in HER2E (*BAG5*). In LumB, 664 genes were differentially expressed (FDR p<0.05), however with typically small fold-changes for the majority of genes. GSEA analysis of this gene set identified only three KEGG pathways that appeared unrelated (**Figure 3B**, **Supplementary Figure S4, Supplementary Table S2**). DEGs were also compared to a list of 219 DNA repair associated genes and three HRD associated gene lists (Peng et al., n=230 [33], and Jacobson et al., n=228 and n=26 [31], listed in **Supplementary Table S3**). Notably, within the 3,400 DEGs identified in the all tumor group comparison, 24-39% of genes in the four gene lists were represented; however, in the specific LumB group comparison with 664 DEGs, the gene representation declined substantially (2-8%) (**Figure 3B, Supplementary Table S2**).

We computed rank scores for eight gene sets (or metagenes) representative of different biological processes (stroma, immune response, lipid, mitotic checkpoint, mitotic progression, basal expression, steroid response, and early response genes) [27] for each tumor to elucidate differences in large scale transcriptional profiles between HRD and HR-proficient tumors. In addition, an RNA-sequencing based Risk of Recurrence (ROR) score, developed as part of the Prosigna prognostic BC assay [34], was also computed as described [28]. In LumB, HRD tumors were associated with higher rank scores for immune response metagenes and proliferation-associated metagenes connected to mitotic progression, while showing lower rank scores for the steroid response metagene consistent with lower levels of ER and PR-stained tumor cells (**Figure 3C**). Higher tumor proliferation observed in HRD compared to HR-proficient LumB tumors was reinforced by both higher ROR scores and Ki67 IHC indices (**Figure 3D**, **Supplementary Figure S4**). In PAM50 Basal tumors, HRD tumors showed higher immune response metagene rank scores compared to HR-proficient tumors, but there was no difference in proliferation or steroid response metagenes (**Figure 3C**). In the HER2E subtype, HRD tumors showed a trend towards higher proliferation, and lower expression of basal and lipid metagenes (**Figure 3C**). Overall, HRD status in ERpHER2n disease is not represented by a unique transcriptional profile. HRD tumors exhibit diverse transcriptional profiles with evidence of higher tumor proliferation and/or immunologic potential in some, but not all PAM50 subtypes.

### Specific immune features of HR-deficient ERpHER2n BC

Although immune infiltration estimates based on RNA-sequencing should ideally be validated *in situ*, for instance through estimation of tumor infiltrating lymphocytes (TILs), previous findings in SCAN-B TNBC tumors have demonstrated that the specific immune expression metagene used above shows high correlation to whole slide hematoxylin and eosin (H&E) TIL counts based on pathologist scoring [35]. To further investigate immune response differences between HRD vs HR-proficient tumors, we analyzed programmed cell death-ligand 1 (PD-L1) mRNA expression (*CD274*) and immune cell types based on CIBERSORTx-deconvoluted RNA-sequencing data. Higher *PD-L1* expression was observed in mainly Basal and LumB HRD tumors, consistent with the immune metagene patterns (**Figure 3E**). In LumB tumors (the subtype with most cases), B-cells were the only significant CIBERSORTx cell type (of B-cells, CD8 T-cells, CD4 T-cells, NK-cells, monocytes, and neutrophils) associated with HR-status (**Figure 3F**). We also analyzed associations of T-cell/B-cell receptor diversity based on computed RNA-sequencing TCR/BCR scores with HRD status. Focusing on the T-cell receptor genes *TRA* and *TRB* (due to read count levels), we observed a higher Shannon-Wiener diversity in HRD tumors compared to HR-proficient tumors for both T-cell receptor genes (**Figure 3G**), consistent with higher immune metagene scores. No differences were observed for the analyzed B-cell receptor genes *IGH*, *IGK*, and *IGL* (Wilcoxon’s test p>0.05).

Finally, we also analyzed Human Leukocyte Antigen (HLA) status based on DNA copy number levels using HLA*LA-HLA typing [36]. HLA*LA-HLA provided data on 13 different loci and 191 unique alleles across the 502 tumors. There was no locus with statistically different proportions (defined as present/absent) between HRD and HR-proficient tumors. While four unique alleles showed different proportions of present/absent between HRD and HR-proficient tumors (Chi-square test p<0.05, example shown in **Figure 3H**), these differences were not significant after adjusting for multiple testing (FDR p>0.05). We also compared specific allele status versus the immune expression metagene rank scores, finding only three unique alleles with a Wilcoxon’s p<0.05 that did not remain significant after adjustment for multiple testing (FDR>0.05, example in **Figure 3I**).

### Genomic features of HR-deficient ERpHER2n BC

We next contrasted genomic features of HRD and HR-proficient tumors within PAM50 subtypes. A variety of features were evaluated including genome-wide patterns of copy number alterations (CNAs), the fraction of the genome altered by CNAs and loss of heterozygosity (LOH), tumor mutational load, tumor ploidy, mutational and rearrangement signatures (**Supplementary Figure S5**). HRD tumors showed higher total mutational load and genomic instability, distributed through the genome (single base substitutions (SBSs), indels, and/or structural variants) compared to HR-proficient tumors, irrespective of PAM50 subtype. HRD tumors did not have statistically different tumor ploidy compared to HR-proficient cases (Wilcoxon’s test p>0.05) but did show more regions of copy number gain or loss and generalized LOH than HR-proficient tumors in the total cohort, LumB, and HER2E tumors (as exemplified for copy number alterations in LumB tumors in **Figure 4A** and furthermore in **Supplementary Figure S5**). We also compared proportions of 25 copy number signatures [30] in tumors stratified by HRDetect and PAM50 status (**Supplementary Figure S5**). Similar to the subtype specific mRNA analyses, this analysis is also statically limited by lower sample numbers in Basal, HER2E, and Normal tumors. In agreement with an HRD phenotype, elevated proportions of CN17 (associated with HRD [30]) were observed in HRD Basal and LumB tumors, while several other statistically significant differences in LumB tumors (like CN4, CN6, CN16, and CN25) appeared outlier driven (**Figure 4B-C**, FDR adjusted Student t-test p<0.05). Combined, these analyses demonstrate that HRD ERpHER2n tumors typically have more complex genomes compared to HR-proficient tumors across PAM50 subtypes.

**Figure 4.**
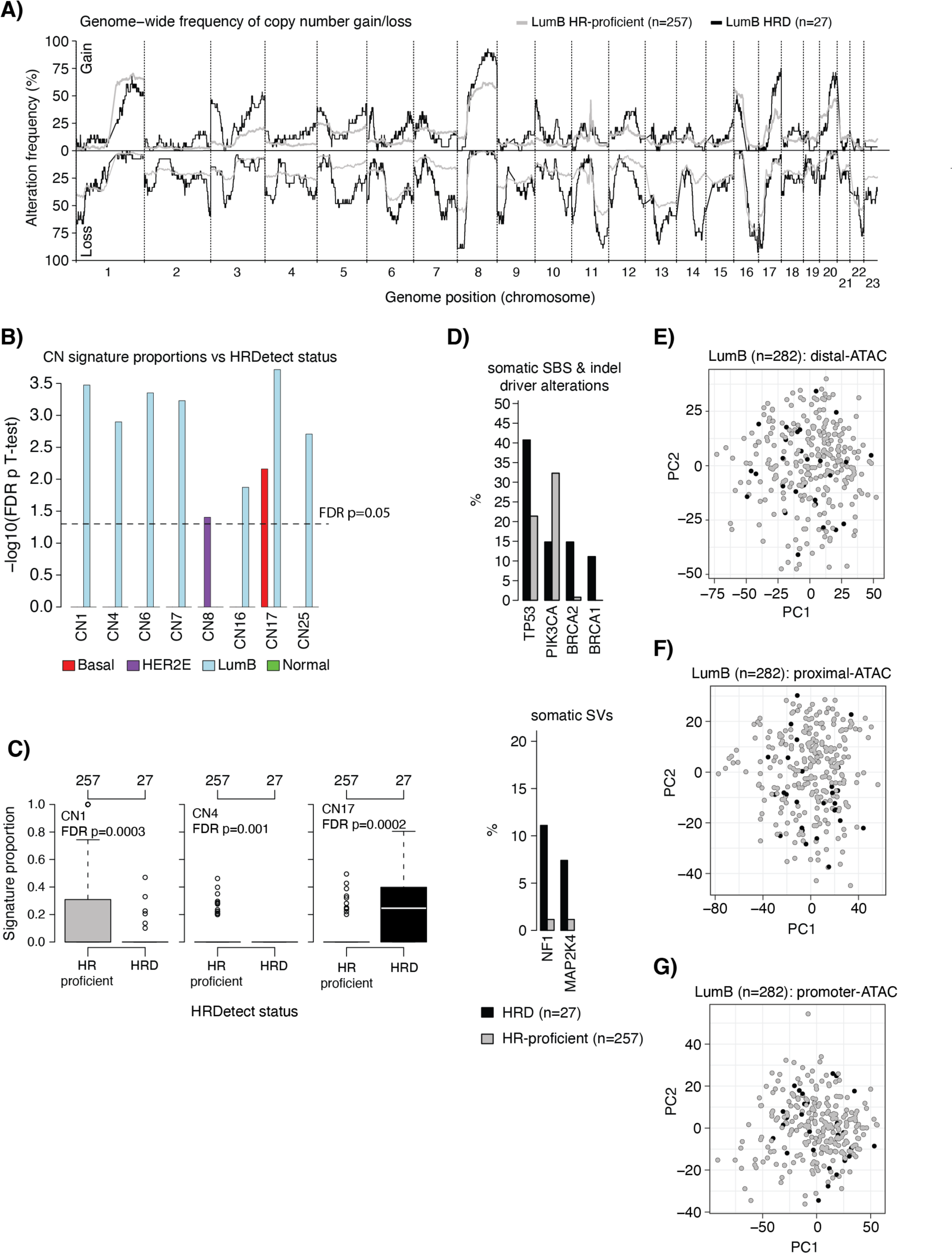
DNA alterations and DNA methylation patterns in ERpHER2n BC with respect to HRD status. (**A**) Genome wide frequency of copy number gain and loss in LumB tumors stratified by HRDetect status. (**B**) Bar plot of FDR adjusted Student T-test values (-log10 transformed) for eight copy number signatures with an FDR adjusted p<0.05 between HR-proficient and HRD tumors stratified by PAM50 subtype. E.g., for CN1, a significant p-value was observed only in LumB tumors. (**C**) Proportions of copy number signature (CN) 1, 4, and 17 in LumB tumors stratified by HRD status. P-value calculated using Student T-test and adjusted for FDR. (**D**) Frequency of driver alterations in LumB tumors that differ between HRD and HR-proficient tumors. Only genes with at least two affected cases and a Fisher’s exact test p<0.1 are shown. (**E**) Principal component analysis of tumor purity adjusted DNA methylation data (beta values) based on the 5000 most variant distal-ATAC CpGs in LumB tumors. Black dots represent tumors with an HRDetect HRD classification. First two principal components (PC1 and PC2) shown. (**F**) Same as in E but for CpGs in a proximal-ATAC context. (**G**) Same as in E, but for CpGs in a promoter-ATAC CpG context. Boxplot elements correspond to: i) center line = median, ii) box limits = upper and lower quartiles, iii) whiskers = 1.5x interquartile range. In boxplots, top axes indicate group sizes.

The relative frequency of somatic driver events (SBSs, indels, and structural variants) in HRD and HR-proficient tumors was also investigated (**Supplementary Figure S5**). The relative proportion of *TP53* drivers was higher in the HRD tumors in all subtypes (example for LumB in **Figure 4D**) and lower *PIK3CA* mutation rate in HRD LumB tumors compared to HR-proficient LumB was observed, in line with previous findings (**Figure 4D**) [22, 31].

### Global DNA methylation features of HR-deficient ERpHER2n BC

To investigate if HRD tumors display different global DNA methylation patterns compared to HR-proficient tumors we performed PCA analysis of tumor purity-adjusted CpG beta values (representing conceptually purer tumor methylomes, see [37]) obtained from Illumina EPIC DNA methylation arrays in the total cohort (n=499), and restricted to the PAM50 subtypes Basal (n=16), HER2E (n=31), and LumB (n=282) due to sample numbers. PCA analyses were performed in three different CpG contexts (gene distal, promoter proximal, and promoter) to acknowledge CpG density differences in the genome. Each CpG set was further filtered to only include the 5000 most variant CpGs mapping to proposed regions of open chromatin defined by ATAC-sequencing (obtained from [38], distal-ATAC, proximal-ATAC, and promoter-ATAC), as this was recently shown in TNBC to define more information rich CpG subsets [39]. Notably, in all PCA comparisons, HRD status did not appear as a distinct divider of variation (**Figure 4E-G** showing LumB and **Supplementary Figure S5** for all results). Next, we performed differential DNA methylation analyses between HRD and HR-proficient tumors in the same sample groups for 741144 CpGs. Only in all tumors and LumB tumors were significantly differentially methylated CpGs identified, but in very small numbers after multiple testing adjustment (n=55 and 4, respectively, Wilcoxon’s test, Bonferroni adjusted p<0.05 and mean absolute difference in beta>0.25 between groups). Together, the unsupervised and supervised DNA methylation analyses suggest that HRD status is not associated with specific or distinct global DNA methylation patterns in ERpHER2n tumors.

### Mechanisms of HRD in ERpHER2n BC

To understand gene inactivation mechanisms underpinning the 42 HRD ERpHER2n tumors, we examined known germline status, somatic variants, and DNA methylation status of promoter CpGs for 219 DNA repair deficiency-associated genes (listed in **Supplementary Table S3**). Thirty (71.4%) cases had potentially causative genetic/epigenetic events involving *BRCA1*, *BRCA2*, *RAD51C*, or *PALB2*, while 28.6% (12/42) of cases lacked a causally implicated event (**Figure 1A**, **Supplementary Table S4**). Among those with a potentially causative event, promoter hypermethylation of *BRCA1*/*RAD51C* accounted for ∼47%, WGS-based somatic alterations (SNVs, indels, structural rearrangements, and homozygous deletions) underpinned ∼43%, and pathogenic germline variants explained 10% of HRD cases. Importantly, clinical germline screening data were not available for all HRD patients in this study due to privacy restrictions, preventing full review of germline data in SCAN-B patients. Consequently, the impact of pathogenic germline variants as an inactivation mechanism is likely underestimated in this cohort.

*BRCA1* and *RAD51C* promoter hypermethylation were observed in 21.4% (9/42) and 11.9% (5/42) of HRD tumors, respectively (**Figure 5A**), with biallelic inactivation through LOH or a somatic variant observed in 92.8% (13/14) of these hypermethylated cases. Promoter hypermethylation of *BRCA1* and *RAD51C* was also frequent within PAM50 subtypes, with 66.7% *BRCA1* promoter methylation frequency alone in PAM50 Basal HRD tumors, 50% frequency of combined *BRCA1* and *RAD51C* hypermethylation in HER2E HRD tumors, and 22% combined frequency in PAM50 LumB HRD tumors (**Figure 5B-D**). We have previously shown that TNBC tumors with inactivated *RAD51C* and *PALB2* present a genetic phenotype similar to *BRCA2*-deficient tumors [9]. To examine whether the type of inactivation mechanism was associated with distinct transcriptional patterns in HRD tumors we performed PCA analysis for all 42 HRD tumors and for the 27 LumB HRD tumors specifically (due to numbers), however not finding that, e.g., tumors with *BRCA2*/*PALB2*/*RAD51C* alterations appeared largely different from *BRCA1* inactivated cases (**Figure 5E**). Substantiating this finding, we reached a similar conclusion regarding inactivation mechanism and global DNA methylation variation using PCA analysis performed as above in LumB HRD tumors specifically (**Supplementary Figure S5**).

**Figure 5.**
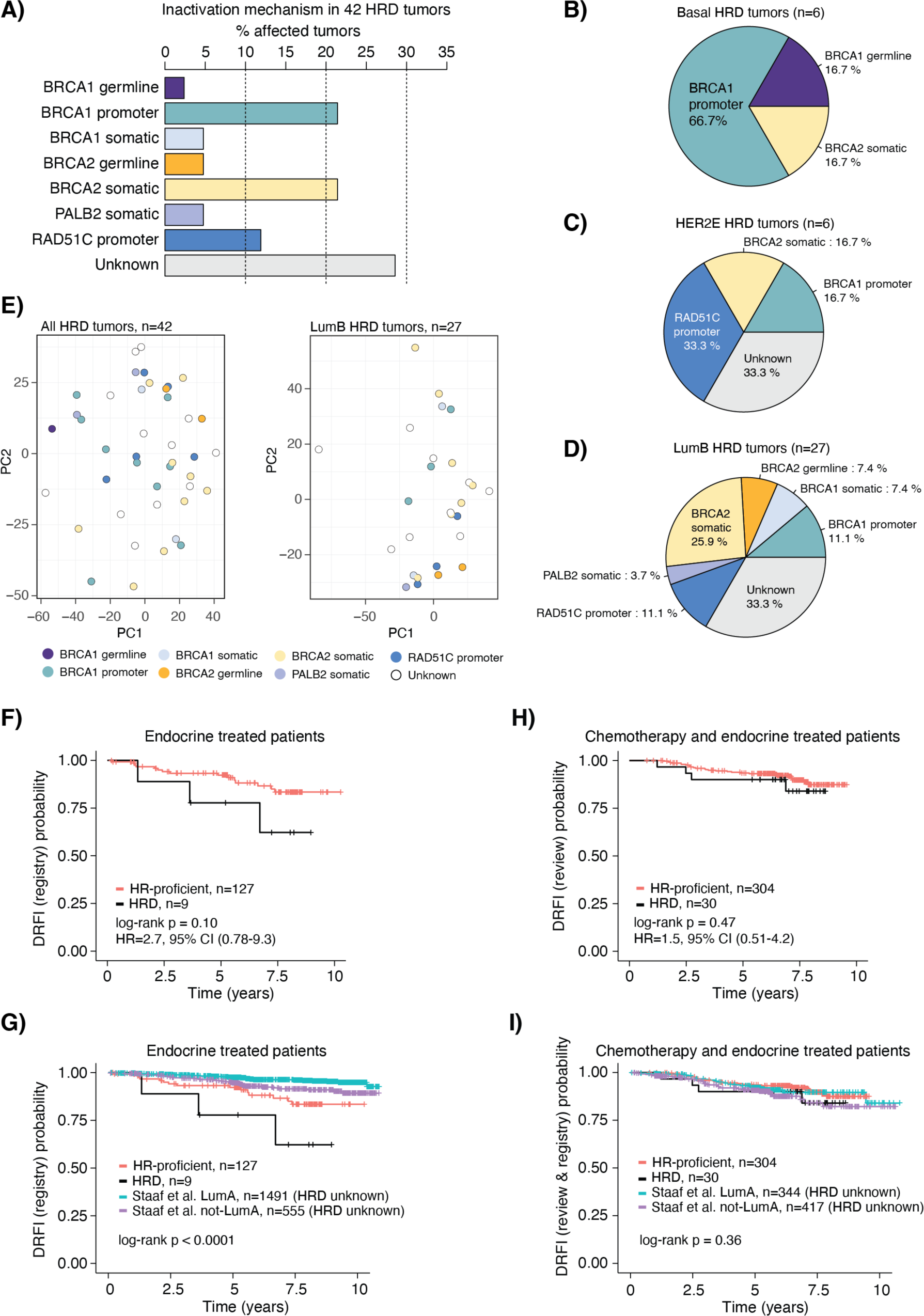
Inactivation mechanisms in HRD tumors and associations with patient outcome. (**A**) All 42 HRD tumors. (**B**) Six HRD PAM50 Basal tumors. (**C**) Six HRD PAM50 HER2E tumors. (**D**) 27 HRD PAM50 LumB tumors. In panels A-D, a few cases exist harboring both promoter methylation and a somatic variant for a gene. For these cases only one of the alterations has been counted as detailed in **Supplementary Table S4**, based, e.g., on correlation to mRNA expression. (**E**) Principal component analysis in all HRD tumors (left) and in LumB HRD tumors (right) using the 5000 most variant genes based on FPKM data for respective group colored by the proposed HRD inactivation mechanism. Principal component 1 and 2 (PC1 and PC2, respectively) are shown. The LumA, Basal, and HER2E groups are excluded due to few cases. (**F**) Kaplan-Meier plot of the association of HRD status with DRFI for patients with HRD and HR-proficient WGS analyzed tumors treated with Endo. DRFI was based on cancer registry data from [28]. Univariate Cox regression hazard ratio and 95% confidence interval shown. (**G**) Kaplan-Meier plot of the association of HRD status with DRFI for HRD and HR-proficient WGS analyzed tumors with inclusion of 2046 additional non-overlapping Endo treated patients with unknown HRD status from [28] stratified by their PAM50 LumA status (LumA or not-LumA). DRFI was based on cancer registry data from [28]. (**H**) Kaplan-Meier plot of the association of HRD status with DRFI for patients with HRD and HR-proficient WGS analyzed tumors treated with ChemoEndo. Clinical review DRFI data was used as endpoint. Univariate Cox regression hazard ratio and 95% confidence interval shown. (**I**) Kaplan-Meier plot of the association of HRD status with DRFI for HRD and HR-proficient WGS analyzed tumors with inclusion of 761 non-overlapping ChemoEndo treated patients with unknown HRD status from [28] stratified by their PAM50 LumA status (LumA or not-LumA). For the 761 patients the DRFI data based on cancer registry data from [28], while for the WGS analyzed samples clinical review data was used as endpoint. In Kaplan-Meier plots the p-value was calculated using the log-rank test.

Regarding other DNA repair deficiency genes, two patients had clinically reported pathogenic germline variants in *ATM*, and one had a pathogenic germline variant in *CHEK2*. However, all tumors showed an intact alternative allele and were HRD-negative. No tumor showed promoter hypermethylation of *ATM* or *CHEK2*. Two HR-proficient tumors had somatic *CHEK2* variants, while seven tumors had somatic *ATM* SBS or indel drivers. Only one of these tumors was HRD (without any variants in *BRCA1*, *BRCA2*, *PALB2*, or *RAD51C*). Together, these data do not support *ATM* or *CHEK2* alterations as important mechanisms for HRD in ERpHER2n BC.

### Association of HRD with prognosis in ERpHER2n BC

Finally, to understand prognostic value of HR-status in ERpHER2n cancers treated with Endo and ChemoEndo, we assessed distant recurrence-free interval (DRFI). In the non-representative Endo WGS cohort (n=136), HRD status was borderline non-significantly associated with differences in DRFI (log-rank p=0.10, **Figure 5F**), although it needs to be stressed that the cohort and the number of HRD cases are small. Because of these limitations, we included an independent non-overlapping cohort of 2,046 Endo-only patients with unknown HRD status [28] to contrast outcome patterns. Notably, the HRD group showed the poorest outcome of all groups despite its sample size, and LumA cases exhibited the best outcome (**Figure 5G**). Supporting the skewness of our HR-proficient WGS Endo cases these patients had a poorer DRFI compared to patients with non-LumA tumors in the independent cohort (log-rank p<0.0001). Thus, although not population-representative and hampered by low numbers, we observe a trend that warrants further consideration – patients with ERpHER2n HRD tumors are unlikely to have good outcomes on Endo treatment alone and may benefit from additional or alternative therapeutic strategies.

In the population-representative ChemoEndo cohort, HR-status was not associated with differences in DRFI (log-rank p=0.47, **Figure 5H**), although a non-significant trend towards poorer overall survival was observed in HRD patients (log-rank p=0.08). Merging data from the WGS ChemoEndo patients with 761 independent, non-overlapping ChemoEndo treated patients with unknown HRD status [28] stratified by PAM50 LumA status demonstrated that all subgroups had approximately similar outcome (**Figure 5I**). Substantiating the latter, in a multivariate Cox regression model including age, lymph node status and tumor grade as covariates, only lymph node status (N0/N+) was statistically significant for DRFI in the combined ChemoEndo treated patient cohort (**Supplementary Figure S6**). Finally, it should be acknowledged that due to the retrospective nature of our cohort with long-term follow-up, no patient received adjuvant therapy including CDK4/6 inhibitors, platinum-based agents, or PARP-inhibitors due to national treatment guidelines at the time.

## DISCUSSION

In this study, we show that HRD tumors occur in ERpHER2n BC, though at a lower frequency than in TNBC. Adjusting for population frequencies of different BC subtypes, we provide the estimate that roughly 1 in 20 ERpHER2n BC and 1 in 9 of all BC are HRD. While a definitive study of HRD frequency in population-representative HER2-positive disease and LumA tumors is warranted, these are unlikely to drastically shift overall estimates suggested in this study as shown in **Figure 1B** (frequency estimates with 20% error spans). Notably, while the SCAN-B study in general has been shown to be population representative [25, 28], it is representative of a specific demographic context of patients (Western Europe/Nordics). Consistently, it should be acknowledged that HRD patterns in ERpHER2n patients may differ in other ethnic contexts. Exemplifying the latter, Feng et al. reported an overall HRD frequency of 34.7% in a smaller study based on Chinese BC patients of unclear population representativity, and nearly three times higher HRD frequency in LumA and LumB patients [23].

From an operational point, this work highlights how global WGS, transcriptomics, and methylation studies can be performed on BC samples collected through routine diagnostic settings across district hospitals in Sweden since 2010 and stored in RNAlater. While successfully achieved previously on 254 TNBC samples [9], this larger study reinforces that snap frozen tissue collections are not necessary for WGS and/or other global multiomic investigations. The current study also provides an overview of the agreement of both DNA and RNA-based HRD classification methods in ERpHER2n BC, illustrating the challenges faced using different approaches when applied to tumor specimens collected in a routine diagnostic setting without specific tumor tissue enrichment by e.g. macro dissection.

Crucially, ERpHER2n HRD tumors occur in all PAM50 subtypes, albeit at different frequencies, and do not present a unique transcriptional or DNA methylation profile, especially when acknowledging underlying molecular subtypes (a concept not typically employed in previous studies). Thus, while a PAM50 Basal subtype would indicate a higher probability for HRD, and a LumA subtype of the opposite, PAM50 subtypes would still not be particularly useful for selecting patients for HRD analysis. ERpHER2n HRD tumors have general but not universal transcriptional characteristics, including enrichment of proliferation markers and immunogenic potential: while in need of *in situ* confirmation, HRD ERpHER2n PAM50 Basal and LumB tumors appear more immune infiltrated, and thus possibly more immunogenic than HR-proficient counterparts. A notable observation in this study was the high HRD frequency (37.5%) in the small subgroup of ERpHER2n tumors subtyped as PAM50 Basal. In these HRD tumors, *BRCA1* gene inactivation occurred in 83.4%, with epigenetic silencing of *BRCA1* by promoter hypermethylation as a dominating feature, similar to TNBC [9]. In contrast, a broader spectrum of inactivated HR genes was observed in LumB tumors, including *RAD51C* and *BRCA1* promoter hypermethylation, but not *BRCA2* and *PALB2* hypermethylation. In HRD ERpHER2n LumB tumors, the *BRCA2*/*PALB2*/*RAD51C* genetic phenotype (see [9]) constitutes 48.1% of tumors, representing a notable difference to TNBC. Still, the key genetic drivers of HRD in ERpHER2n disease appear similar to TNBC [9], i.e., *BRCA1*, *BRCA2*, *PALB2*, and *RAD51C*. Together, our observations support epigenetic silencing as a major HRD cause in both ERpHER2n BC and TNBC [9]. Overall, in this study a putative inactivation mechanism could be assigned to 71.4% of HRD-positive tumors, representing a conservative lower estimate given the restricted availability of germline status for breast cancer susceptibility genes in our cohort.

A key objective of this study was to investigate prognostic associations of HRD with conventional standard-of-care treatment groups. In our WGS-analyzed Endo cohort, the small collection of HRD tumors stood out as a potential poor outcome subgroup, consistent with their generally lower steroid response and higher tumor proliferation scores compared to HR-proficient tumors. However, the skewing of the Endo cohort towards a more aggressive phenotype emphasizes the need for additional studies assessing the prognostic relevance of HRD in Endo only patients given the limited size of the current cohort. Still, these analyses suggest that patients with HRD ERpHER2n tumors may be at risk of poor outcome if left on Endo treatment alone and should be considered for additional systemic therapy. Population-representative ChemoEndo-treated patients with HRD tumors did not show different DRFI compared to patients with HR-proficient tumors, nor indeed to other stratification statuses including PAM50 LumA. This key, negative finding, opposite to our TNBC study [9] stresses the need to continue the search for biomarkers that can stratify ERpHER2n ChemoEndo patients into refined outcome groups to further tailor adjuvant therapies and treatment (de)escalation options. As SCAN-B is not a randomized clinical trial but a representative view of real-world oncology practice, it should be stressed that patient selection for adjuvant therapy (Endo or ChemoEndo) and other specific treatment regimens is not necessarily the same today as during the 2010-2014 inclusion period. For instance, due to national treatment guidelines at the time, no patient received adjuvant therapy including CDK4/6 inhibitors, platinum-based agents or PARP-inhibitors, requiring additional studies to determine the actual prognostic relevance of HRD (irrespective of inactivation mechanism) in these treatment contexts, as the predictive value of HRD is conflicting for, e.g., platinum-based agents [40]. Still, while our patients are not treated according to the latest recommended guidelines, it should be acknowledged that such molecularly profiled SOC cohorts with long-term follow-up are currently simply not available. While there is a lack of consensus regarding how best to manage HRD ERpHER2n BCs, arguments in favor of considering alternative systemic therapies are two-fold. First, HRD has been associated with more favorable patient outcome after adjuvant chemotherapy in primary TNBC [9]. Second, while in need of substantiation, the small GeparOLA study within early HER2-negative HRD BC reported that patients’ hormone receptor-positive HRD tumors exhibited higher response rates to a combination treatment including olaparib [41, 42]. Whether ERpHER2n *BRCA1*/*BRCA2* germline or HRD patients will respond to CDK4/6 inhibitors or antibody-drug conjugates (ADCs) remains unclear as clinical trial data are scarce [43]. Regardless, while this study raises the hypothesis that patients diagnosed with ERpHER2n HRD tumors may benefit from adjuvant ChemoEndo compared to patients diagnosed with HR-proficient tumors, this needs to be proven in randomized clinical trials.

Our demonstration of increased immune response signatures in HRD ERpHER2n tumors based on RNA-sequencing data is interesting and in line with previous observations [22] given ongoing clinical trials involving checkpoint inhibitors in patients with ERpHER2n disease. While RNA-sequencing immune patterns should be confirmed *in situ* by for example TIL estimation, we recently demonstrated in TNBC that the specific RNA-sequencing immune metagene correlated well with whole slide H&E pathology estimated TIL counts [35]. Immune checkpoint inhibitors are currently being explored for early hormone receptor-positive HER2-negative breast cancer, e.g., in the KEYNOTE-756 and CheckMate 7FL trials [44, 45]. The CheckMate 7FL trial demonstrated that adding the anti-PD-1 agent nivolumab to neoadjuvant chemotherapy significantly improved pathological complete response rates in high-risk ERpHER2n breast cancer, particularly in tumors with high stromal TIL levels and/or PD-L1 positivity. Moreover, the investigators observed a greater benefit in patients with low ER expressing tumors (<10%), where these features of immunogenicity have previously been shown to be elevated [44, 46]. Although ERpHER2n HRD cases in this study were defined by an ER expression ≥10%, our study indicates increased *PD-L1* mRNA expression and immune activation by mRNA signatures in ERpHER2n HRD patients, suggesting that immunotherapy could be of potential clinical benefit to this small patient subset. Finally, in the GIADA trial it was reported that the combination of a basal-like intrinsic subtype (herein shown to display a high HRD frequence) and high TILs could predict pathological complete response after neoadjuvant treatment with chemotherapy, immune checkpoint inhibition, and endocrine therapy in premenopausal women with aggressive hormone receptor-positive (ER and/or PR ≥10%) / HER2-negative breast cancer [47]. Together, while several of these features match characteristics of HRD tumors, it remains to be fully determined whether HRD tumors would benefit from this therapeutic scenario.

Taken together, our study highlights the HRD phenotype in ERpHER2n tumors as a marker of aggressiveness, even when compared to HR-proficient tumors within the same molecular subtype, and an interesting association with elevated immune response that should be pursued in more depth. We demonstrate that genomic HRD provides additional information beyond intrinsic molecular BC subtypes in categorizing tumors and understanding which patients might need further treatment. Moreover, this study shows that HRD is much less frequent in ERpHER2n tumors compared to TNBC, suggesting that the overall HRD frequency in a Nordic/Western European demographic context is likely between 10-13%, but that the same key drivers of DNA repair deficiency are identified in both breast cancer groups.

## METHODS

### SCAN-B ERpHER2n breast cancer cohort

Patients with primary ERpHER2n BC were enrolled in the SCAN-B study (NCT02306096) during 2010-2014. Ethical approval was given for the SCAN-B study (Registration numbers 2009/658, 2010/383, 2012/58, 2013/459, 2014/521, 2015/277, 2016/541, 2016/742, 2016/944, 2018/267, 2019/01252, and 2024-02040-02) by the Regional Ethical Review Board in Lund, Sweden, governed by the Swedish Ethical Review Authority, Box 2110, 750 02 Uppsala, Sweden. All patients provided written informed consent prior to enrolment. All analyses were performed in accordance with patient consent and ethical regulations and decisions. The final patient cohort comprised 502 patients with successful WGS (see CONSORT diagram in **Supplementary Figure S1A, Supplementary Methods**, **Table 1** and **Supplementary Table S1** for patient characteristics). Of the 502 patients, 138 received endocrine adjuvant therapy (Endo) forming the final Endo treated group for survival analysis. Of these 138 patients, all had available OS data, while 136 patients had available DRFI data from [28]. Of remaining patients, 352 received combined adjuvant chemotherapy and endocrine therapy (ChemoEndo) based on registry data [28], where 339 of 352 (96.3%) cases were confirmed by clinical review to have received chemotherapy, forming the final ChemoEndo group for survival analysis. The main chemotherapeutic strategy was fluorouracil+epirubicin hydrochloride+cyclophosphamide (FEC) combined with a taxane (typically docetaxel) (92.6% of the 339 ChemoEndo patients). In the SCAN-B cohort, ER and PR data are based on routine clinical IHC analysis, while HER2-status was determined from routine clinical IHC and FISH analysis as reported in the Swedish national breast cancer quality registry. Specifically, ER-positivity was defined as ≥10% of tumor cells being IHC-stained according to current Swedish national guidelines. Fresh tumor tissue from SCAN-B patients were preserved in RNAlater™ Stabilization Solution (Invitrogen) as outlined [25].

### Non-related ERpHER2n gene expression SCAN-B cohort

Based on our recent study, 4,924 tumors ERpHER2n tumors, including all but five WGS cases due to lack of lymph node registry data, were identified providing proportions of PAM50 subtypes in ERpHER2n disease [28]. Of these, 4,427 did not overlap with the WGS cohort. From the set of 4,427 patients, PAM50 nearest subtype classification, clinical data, and DRFI data were available for 2,046 Endo-only treated patients and 761 ChemoEndo treated patients. For these patients, HRD status is unknown, while RNA-sequencing and PAM50 subtyping were performed identical to the WGS analyzed cohort. Clinical data and patient outcome data for these patients are based on cancer registry data from [28] (see **Supplementary Methods** for additional information).

### HRD estimate in HER2-positive BC

In the study by Nik-Zainal et al., 73 HER2-amplified tumors were analyzed by WGS, with three cases being called HRD (4.1%) by HRDetect [6].

### Gene expression analyses

RNA-sequencing data were available as fragments per kilobase million (FPKM) values from [28] for all patients, including also PAM50 molecular subtypes and ROR risk groups, both based on nearest centroid classifications. Gene expression analyses including molecular subtyping, ROR classification, biological metagenes, supervised and unsupervised analyses, immune cell type deconvolution, T-cell receptor (TCR) and B-cell receptor (BCR) repertoire analysis, and functional enrichment analyses are further detailed in **Supplementary Methods**. Gene expression-based HRD classification was performed using the 228 gene HRD nearest centroid classifier (comprising one HRD centroid and one HR-proficient centroid) reported by Jacobson et al. [31]. Prior to classification using FPKM data, an offset of 0.1 was added, the data was log2-transformed and mean-centered. For each sample, the Pearson correlation to each centroid was computed and also an HRD score as outlined by Jacobson et al. Classification was performed using either the highest correlation, or specific correlation cut-offs applied to the HRD centroid values.

### Whole genome sequencing analysis and HRD prediction

Paired tumor and normal blood cells WGS were performed at Novogene (Novogene, UK) using 150 bp paired-end sequencing to reach 120-150 GB of sequence, resulting in an effective median tumor coverage of 36x in the final cohort of sequenced tumors. Alignment was performed versus GRCh38. WGS pre-processing, CNA analysis, mutational (Caveman v1.13.15), indel (Pindel v3.2.0), and rearrangement signature analysis (BRASS) were performed as detailed in **Supplementary Methods**. HRD status (HRD-positive/HRD-negative) was called with HRDetect [8], using ≥0.9 as the probability threshold for calling HRD (HRDetect-high), <0.1 as HRDetect-low, and 0.1-0.9 as HRDetect-intermediate. In analyses involving a two-group comparison, HRDetect-high tumors were considered HR-deficient (HRD), while HRDetect-low and HRDetect-intermediate tumors were combined into a single HR-proficient group. Copy number signature proportions per tumor for 25 signatures defined by Steele et al. [30] were calculated using segmented ASCAT data and code available from COSMIC v3.4 (https://cancer.sanger.ac.uk/signatures/cn/). scarHRD HRD scores were calculated based on segmented ASCAT data using R scripts available from the project’s GitHub [29]. Tumors with summarized HRD scores >42 were called as HRD, otherwise HR-proficient. CHORD [24] HRD classification was performed by first combining the Caveman (SNVs) and Pindel (indel) VCF files into one common SNV VCF file per sample with Bcftools “concat” with option “-a” [48]. After the merge, only variants tagged PASS were retained. The structural variants (SVs) BRASS VCF files were used as is. HR deficiency was predicted with CHORD in single sample mode using the merged SNV and BRASS SV files as input for each sample. Reference genome was GRCh38 and the option “-include_non_pass” was used to retain all variants in the BRASS VCF in CHORD prediction (BRASS does not tag variants as PASS/noPASS and the PASS tag is required by CHORD). HLA analysis was performed using ASCAT data and the HLA*LA-HLA typing software [36]. Detected alleles were filtered as outlined in **Supplementary Methods**.

### DNA methylation analysis

DNA methylation profiling of 499 of 502 tumors, including all 42 HRD-positive tumors, was performed using Illumina Infinium EPIC v1 beadarrays according to manufacturer’s instructions as reported by Hohmann et al. [49]. DNA methylation analysis was performed on the same DNA extracts as used for WGS. Methylation profiling was performed by the SNP&SEQ Technology Platform in Uppsala (www.genotyping.se). Beta values, representing the level of methylation, were computed in a sample-by-sample context using the minfi R package v1.44 function *preprocessNoob()*, Infinium probe normalized using the approach described in [50], and filtered according to Aine et al. [39] leaving 741144 CpGs for final analysis. DNA methylation profiles are available as GSE278586 through Gene Expression Omnibus. Adjustment of beta values for tumor purity using the PureBeta pipeline [51] (referred to as tumor purity-adjusted beta values) was performed as outlined in **Supplementary Methods**. Annotation of CpGs was performed as outlined by Aine et al. [39] and is, together with details on unsupervised and supervised analyses, further described in **Supplementary Methods**.

### Survival analyses

Survival analyses were performed in the final Endo and ChemoEndo treatment groups. For the Endo group, cancer registry outcome data from [28] were used (due to incomplete available clinical review of all cases), whereas for the ChemoEndo group clinical review data were available for the 339 patients. Distant recurrence-free interval (DRFI), defined according to the STEEP criteria [52], and overall survival (OS) were used as primary endpoints for both treatment groups. Median outcome measures in censored patients were equal to 7.2 and 8.6 years for DRFI and OS, respectively, in the ChemoEndo cohort, and 6.5 and 8.4 years for DRFI and OS, respectively, in the Endo cohort. Survival analyses were performed in R (v4.4.2) using the *survival* (v3.7.0) and *survminer* (v0.5.0) packages. Survival curves were estimated using the Kaplan-Meier method and compared using the log-rank test. Hazard ratios were calculated through univariable and multivariable Cox regression using the *coxph* R function.

## Data Availability

DNA methylation data used in this study is accessible from Gene Expression Omnibus.
RNA-sequencing data used in this study is available from an open repository associated with a previously reported study. Summarized WGS data is available as supplementary data. WGS sequence data is protected by Swedish law as sensitive patient information.

https://www.ncbi.nlm.nih.gov/geo/query/acc.cgi?acc=GSE278586

https://data.mendeley.com/datasets/yzxtxn4nmd/3

## ACKNOWLEDGEMENTS

The authors would like to acknowledge all patients and clinicians participating in the SCAN-B study, personnel at the central SCAN-B laboratory at the Division of Oncology, Department of Clinical Sciences Lund, Lund University, the Swedish national breast cancer quality registry (NKBC), Regional Cancer Center South, RBC Syd, and the South Sweden Breast Cancer Group (SSBCG). Methylation profiling of the SCAN-B cohort was performed by the SNP&SEQ Technology Platform in Uppsala (www.genotyping.se). The facility is part of the National Genomics Infrastructure supported by the Swedish Research Council for Infrastructures and Science for Life Laboratory, Sweden. Financial support for this study was provided by the Swedish Cancer Society (CAN 2021/1407 JS, 2024/3591 JS), the Mrs Berta Kamprad Foundation (FBKS-2024-14 JS), the Swedish Research Council (2021-01800 JS), the Mats Paulsson Foundation (ÅB), the Cancera Foundation (ÅB), the National Society of Breast Cancer Associations in Sweden (JS), and Swedish governmental funding (ALF, grant 2022/0021 JS). The funders had no role in study design, data collection and analysis, decision to publish, or preparation of the paper.

