## Supplementary methods for "Homologous recombination deficiency in primary ER-positive and HER2-negative breast cancer"

### SCAN-B ERpHER2n breast cancer cohort

Patients with primary ERpHER2n BC were enrolled in the SCAN-B study during 2010-2014. Ethical approval was given for the SCAN-B study (Registration numbers 2009/658, 2010/383, 2012/58, 2013/459, 2014/521, 2015/277, 2016/541, 2016/742, 2016/944, 2018/267, 2019/01252, and 2024-02040-02) by the Regional Ethical Review Board in Lund, Sweden, governed by the Swedish Ethical Review Authority, Box 2110, 750 02 Uppsala, Sweden. All patients provided written informed consent prior to enrolment. All analyses were performed in accordance with patient consent and ethical regulations and decisions.

During 2010 to 2014, 3066 patients were diagnosed with primary ERpHER2n breast cancer in the Skåne Healthcare region, Sweden, based on data from the national breast cancer registry. Of these, 2611 were enrolled in the SCAN-B study and 1834 of the 2611 (70.2%) patients were included in the study by Staaf et al. [1], based on availability of RNA-sequencing data (including PAM50 subtype classification as detailed in [1]). Further inclusion and exclusion criteria for the 1834 patients are described in detail in [1], with clinical data provided from the national breast cancer registry available from [1]. ERpHER2n patients in the study by Staaf et al. have been shown to representative of the patient demographics in the total catchment region during the inclusion period (see [1]). From the 1834 patients, 533 were selected for WGS analysis based on a combination of PAM50 subtype classification, administered treatment based on clinical data, availability and sufficient amount of tumor and matched normal DNA for whole genome sequencing (WGS). Patients were prioritized for WGS based on: i) ChemoEndo adjuvant therapy irrespective of PAM50 subtype to allow survival analyses specifically in this treatment group, ii) a PAM50 Basal, Luminal B (LumB), or HER2-enriched (HER2E) subtype irrespective of adjuvant therapy to allow for comparison of HRD frequency in these PAM50 subtypes, and iii) an addition of a small set of Luminal A (LumA) tumors. After WGS quality assessment and sample filtering, 502 patients remained, forming the final analysis cohort. The absolute major cause of WGS failure was estimated low tumor cell content in tumor tissue. Of the 502 patients, 138 (27.5%) received endocrine adjuvant therapy (Endo), while 352 (70.1%) received combined adjuvant chemotherapy and endocrine therapy (ChemoEndo) according to cancer registry data from [1] (remaining cases had other combinations or no treatment indicated in [1]). Based on available clinical chart review data, 339 of 352 (96.3%) ChemoEndo cases were confirmed to be treated with chemotherapy, forming the final ChemoEndo treatment assessment group for survival analysis. From recorded clinical chart review data, the main chemotherapy combination was fluorouracil+epirubicin hydrochloride+cyclophosphamide (FEC) combined with a taxane (typically docetaxel) (92.6% of the 339 ChemoEndo treated patients). The 138 patients receiving adjuvant Endo based on data from [1] formed the final Endo treatment assessment group for survival analysis. Based on data from [2], 57 of the 502 patients had been referred to counseling and genetic screening according to practitioner's choice based on at-the-time current guidelines, a clinical decision that is completely separated from enrollment in the SCAN-B study. Variants had been classified as benign, likely benign, uncertain, likely pathogenic, or pathogenic with clinical significance compared to information on ClinVar[3] at date of accession (May 2020) [2]. Similar to Nacer et al. [2], likely pathogenic and pathogenic variants were combined and referred to only as pathogenic (PV, similar to [4]), and likely benign and benign variants as benign. All genomic analyses were performed on core biopsies or surgical resections prior to adjuvant treatment, and each patient is represented by a single assay for respective method (whole genome sequencing, RNA-sequencing, DNA methylation analysis). A CONSORT diagram is available in Supplementary Figure S1A,

**Non-overlapping SCAN-B ERpHER2n patient cohort**

From the study by Staaf et al. [1] we identified 4,427 patients diagnosed 2010-2018 with ERpHER2n tumors that did not overlap with patients in the WGS cohort. These patients were defined as eligible for follow-up (FU=1), with available clinical data, including lymph node status (LNx) based on data obtained from [1]. To note, all but 5 patients in the WGS cohort represent a subset of patients in [1] defined as eligible for follow-up (FU=1), with available clinical data, including lymph node status (LNx). Therefore, the combined sum of WGS and non-WGS patients fulfilling the above constraints based on data from [1] is 4,427+502-5=4,924. These 4,924 patients were used for definition of population representative PAM50 subtype frequency. RNA-sequencing data was available for both cohorts (WGS and non-overlapping) performed, processed and analyzed the same way. Clinical data and outcome data are based on cancer registry data as reported in [1].

**Statistical tests**

All p-values reported from statistical tests are two-sided if not otherwise specified.

**RNA-sequencing analysis**

RNA-sequencing data were available as fragments per kilobase million (FPKM) values from [1] for all patients (this study outlines also the basic data processing steps). Based on the same publication we also obtained PAM50 molecular subtypes (referred to as NCN subtypes in [1]) and Risk Of Recurrence (ROR scores, column NCN-ROR-T0).

Based on FPKM data, gene expression-based rank scores for eight biological metagenes in breast cancer originally defined by Fredlund et al. [5] (termed basal, lipid, mitotic progression, mitotic checkpoint immune response, early response, steroid response, and stroma) were calculated as described by Nacer et al. [2]. Rank scores were computed individually for each tumor from FPKM data without any further normalization or data centering.

Differential, supervised, gene expression analysis was performed using Student t-test on FPKM data that was (i) offset by +0.1 and (ii) log2 transformed, and FDR adjusted for multiple testing using the *p.adjust* R function. Significant genes were in addition required to have mean group FPKM >1 in both test groups to exclude genes with statistical significance but overall very low expression. Principal component analysis (PCA) was performed using the R function *prcomp*() with the parameters center = TRUE, scale = TRUE, retx = TRUE. In the analysis FPKM data was offset by +0.1 and log2 transformed prior to PCA.

CibersortX estimates of immune cell proportions, based on RNA-sequencing data, were obtained for tumors from Nacer et al. [2], using a p-value cut-off of $p<0.05$ as described.

Pathway analysis was performed using the R ClusterProfiler package (v4.12.6) [6] and the R package implementations of KEGG, Gene Ontology (org.Hs.eg.db, v3.19.1), Reactome (ReactomePA v1.48.0), and Molecular Signatures Database (MsigDB, msigdbr v10.0.1) gene sets associated with Gene Set Enrichment Analysis (GSEA), as outlined in the ClusterProfiler vignette for the respective analysis. An adjusted p-value <0.05 was used as the significance threshold in all analyses. A list including only significant genes was used as input to the analyses. If a gene universe was required, then the full set of 19675 genes for which FPKM data was available was used.

***T cell receptor (TCR) and B cell receptor (BCR) repertoire analysis***

RNAseq fastq files for SCAN-B tumors were processed with *MiXCR* (v4.5.0) using the *rna-seq* preset and specifying species *hsa* [7, 8]. For the divergence analysis, we used *post-analysis* and *individual* presets, enabling the calculation of CDR3 metrics, Shannon-Wiener diversity measurement, and gene segment usage for all tumors as a single group.

**Whole genome sequencing analysis**

For each SCAN-B patient, a tumor sample taken at surgery or as a core biopsy, and matched blood DNA was sequenced at Novogene (Novogene, UK) using 150bp paired end sequencing on an Illumina Novaseq to reach 120-150GB of sequence, resulting in a tumor coverage between 26-50X (median 36X) after duplicate removal and final filtering. Resulting BAM files were aligned to the reference human genome (GRCh38) using dockstore-cgpmap v3.2.0 implementing bwa mem v0.7.17-r1188 https://quay.io/repository/wtsicgp/dockstore-cgpmap. Mutation calling of WGS data for SCAN-B tumors was performed as described previously [9]. Briefly, the mutation calling pipeline was containerized within dockstore-cgpwgs v2.1.1 ( https://quay.io/repository/wtsicgp/dockstore-cgpwgs) , implementing Caveman v1.13.15 for somatic substitution calling (Cancer Variants through Expectation Maximization: http://cancerit.github.io/CaVEMan/), Pindel v3.2.0 for detection of somatic small insertions and deletions (http://cancerit.github.io/cgpPindel/), and BRASS for structural rearrangements (BReakpoint AnalySiS; https://github.com/cancerit/BRASS). ASCAT v4.2.1 (https://github.com/cancerit/ascatNgs) was used to supply average ploidy and purity inputs for Caveman. Additional filtering was applied as follows; single base substitutions (SBSs) were filtered based on a PASS criterion, CLPM=0.00, and ASMD>=140, small indels were filtered by QUAL>=250 & REP<10, and structural variants filtered for BRASS assembly score >0 indicating successful de novo local assembly using Velvet to determine exact coordinates and features of breakpoint junction sequence.

Mutational signatures were assigned using Fit Multi-Step (FitMS) [10] (https://github.com/Nik-Zainal-Group/signature.tools.lib) and the following parameters were used. For substitution signatures, reference SBS previously identified in breast cancer were used in combination with high confidence rare signatures from any organ. A fixed threshold of 5% of the total mutations contributing to the signature was applied before assigning a signature and an error reduction of 20% used. Rare signature assignment was manually reviewed. For structural rearrangement signatures, reference signatures previously identified in breast cancer were fitted to all samples with a total number of rearrangements greater than 25. In addition, mutational signatures were only assigned if a minimum of 5 variants and a minimum percentage of 5% of the total rearrangements were contributing to the signature. The resulting signatures were used as input to HRDetect [11].

Somatic mutations were annotated to Ensembl v91 for using (https://github.com/cancerit/VAGrENT). Non-synonymous point mutations and small indels were assessed for potential driver mutations by comparison to the list of genes in the Cancer Gene Census (https://cancer.sanger.ac.uk/census) and genes previously identified as breast cancer driver genes from a previous study [9]. Mutations within these genes were considered to be potential drivers if the same mutation exists multiple times in the COSMIC database or were reported as pathogenic or likely pathogenic in cancer in ClinVar. In addition, mutations in genes which are reported in the Cancer Gene Census as tumor suppressor genes (excluding those genes where the know mechanism of mutation was restricted to gene fusions) were considered to be potential drivers if the mutation was predicted to result in a premature truncation (nonsense, essential splice, frameshift mutations). Loss of all wild type alleles in tumor suppressor genes due to loss of heterozygosity was assessed using the ASCAT copy number for the corresponding segment.

WGS data was processed for copy number analysis by a modified ASCAT v3.1.2 version. Necessary reference files for this version are available at https://github.com/VanLoo-lab/ascat/tree/master/ReferenceFiles/WGS. Changes to the ASCAT algorithm are outlined at https://github.com/nnordborg/ascat/tree/scanb ("Changes in the SCANB-fork"). Baseline parameters were imbalance.test=bimodality_coefficient and tau=0.4. ASCAT segments were called as copy number gain or loss considering the tumor ploidy as described by Staaf et al. [12].

**HLA analysis**

HLA analysis was performed using ASCAT WGS data and the HLA*LA-HLA typing software [13]. Detected alleles were filtered using the variables perfectG==1, proportionkMersCovered==1.

**DNA methylation analysis**

Matched Illumina EPIC V1 DNA methylation profiles for 499 of 502 tumors were obtained from GSE278586 [14]. Beta values, representing the level of methylation, were computed in a sample-by-sample context using the minfi R package v1.44 function *preprocessNoob()*, Infinium probe normalized using the approach described in [15], and filtered according to Aine et al. [16], leaving 741144 CpGs for final analysis. Promoter methylation status of HR associated genes were assessed based on promoter associated CpGs using manual inspection of normalized beta values.

For unsupervised and supervised analyses we used beta values adjusted for tumor purity as conceptually outlined by [17]. Adjustment of beta values for tumor purity using the PureBeta pipeline [18] (referred to as tumor purity adjusted beta values) were performed using WGS-based tumor purity estimates (ASCAT values) as input values and the previously normalized beta values. We applied the function with default parameters (including the refitting option set to false) and used reference CpG models previously established by Aine et al. [16] in TNBC. Unsupervised analysis using principal component analysis was performed using the R function prcomp() with the parameters center = TRUE, scale = TRUE, retx = TRUE on tumor purity adjusted beta values. For identifying differentially methylated CpGs, tumor purity adjusted beta values were compared using the Wilcoxon test for two group comparisons or the Kruskal-Wallis test if more than two test groups. For each CpG, if the standard deviation in a comparison was 0, then the p-value was set to 1. When filtering CpGs, we also used the absolute difference between mean beta per test group as an additional requirement to avoid obtaining significantly differentially methylated CpGs with very small group differences in beta.

CpGs used in supervised and unsupervised analyses were annotated as outlined by Aine et al. [16]. Briefly, we compiled a custom feature annotation set for each CpG probe on the Illumina EPIC methylation platform using the same methodology described for the Illumina HumanMethylation450K array in Staaf and Aine [17]. This included assigning CpGs to a gene-centric context defined as promoter (+/- 500 bp centered on gene transcription start site, TSS), proximal (+/- 5 kbp centered on TSS and excluding the promoter window), or distal (>5 kbp from TSS) based on their genomic coordinates (referred to as genic context). For the gene-centric annotations, a consensus transcript model based on GENCODE v27 protein coding genes matching SCAN-B RNAseq data was built for each gene by collapsing of exons. The 5' most base was assigned as the consensus TSS and the 3' most base as the consensus transcription termination site. Probes were also assigned to a CpG-centric context defined as CpG island (CGI), shore, or ocean (referred to as CGI context) [19]. Local CpG density metrics (e.g., O/E) and contextual classifications for each probe were obtained using the methods of Saxonov et al. [20] for high (HCG) and low (LCG) CpG content and of Weber et al. [21] for HCP, ICP, and LCP. EPIC probe overlaps with ATAC-seq peak data generated on 74 TCGA breast cancer samples by Corces et al. [22] were calculated and used as a proxy for differentially open chromatin in breast cancer. Additionally, ENCODE candidate cis-regulatory elements

[23] and ENCODE ChIP-seq peak overlaps for 340 transcription factors in 130 cell lines [24] were used to assess the regulatory potential of EPIC CpGs.

Methylation profiling of the SCAN-B cohort was performed by the SNP&SEQ Technology Platform in Uppsala (www.genotyping.se). The facility is part of the National Genomics Infrastructure supported by the Swedish Research Council for Infrastructures and Science for Life Laboratory, Sweden.

**Survival analysis**

Survival analyses were performed in the final Endo and ChemoEndo treatment groups. For the Endo group, cancer registry outcome data from [1] was used (due to incomplete available clinical review of all cases), whereas for the ChemoEndo group clinical review data was available for the 339 patients. Distant recurrence-free interval (DRFI), defined according to the STEEP criteria [25], and overall survival (OS) were used as primary endpoints for both treatment groups. The median outcome measures in censored patients were equal to 7.2 and 8.6 years for DRFI and OS, respectively, in the ChemoEndo cohort, and 6.5 and 8.4 years for DRFI and OS, respectively, in the Endo cohort. Survival analyses were performed in R (v4.2.2) using the *survival* (v3.4.0) and *survminer* (v0.4.9) packages. Survival curves were estimated using the Kaplan-Meier method and compared using the log-rank test.
