## Supplementary Figure S1 for "Homologous recombination deficiency in primary ER-positive and HER2-negative breast cancer"

A)

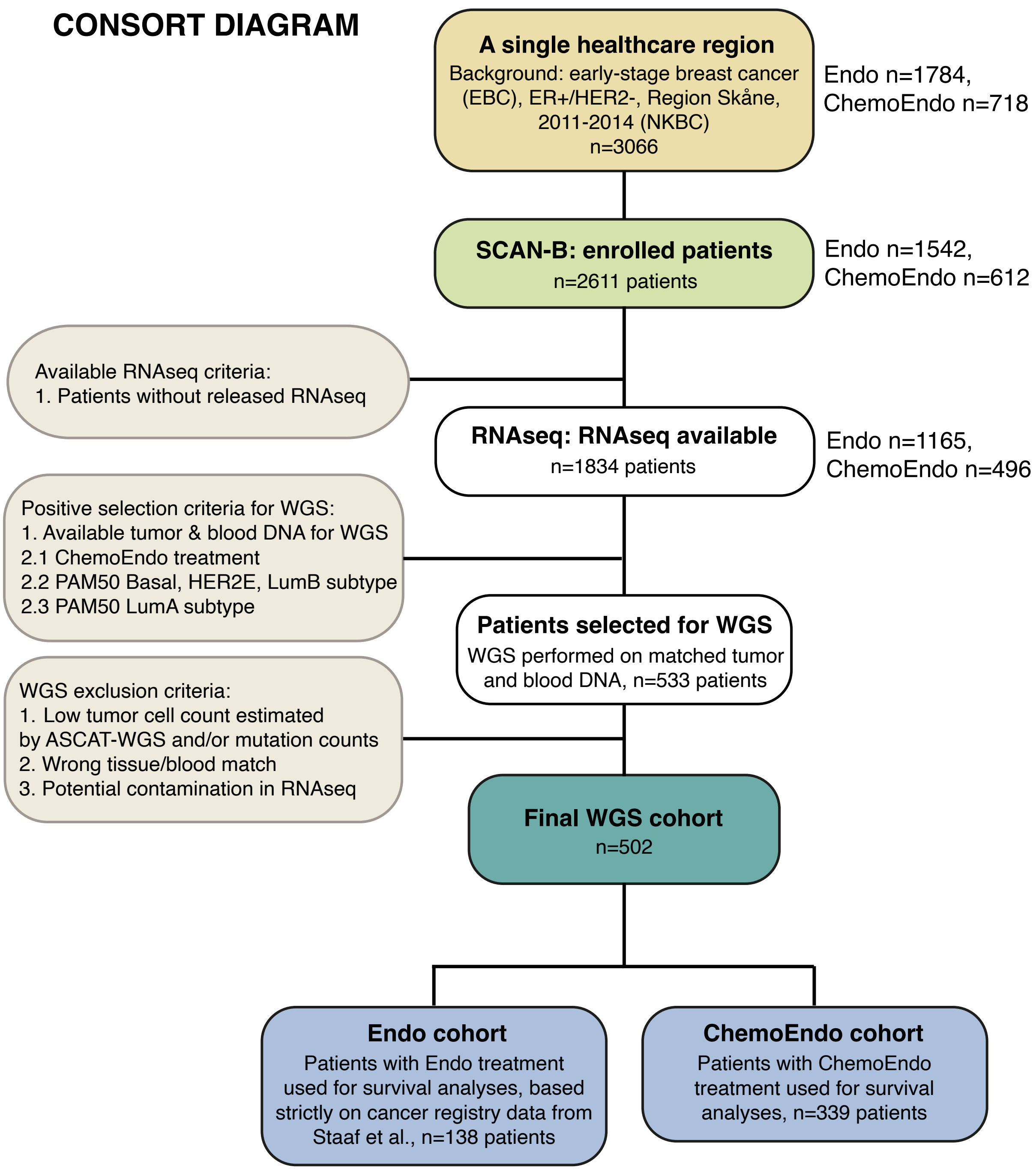


**Supplementary Figure S1. CONSORT and treatment group representativity. (A)** CONSORT diagram. NKBC: Swedish National Quality Registry for Breast cancer. **(B)** Study overview, with key analyses and external cohorts used. **(C)** Representativity of the WGS profiled Endo treatment group compared to background patient populations diagnosed during 2010-2014 for different clinical variables obtained from the NKBC registry in the catchment region (RS: Skane healthcare region). EBC: Early breast cancer (surgically treated). Top axis in barplots corresponds to sample numbers. **(D)** Representativity of the WGS profiled ChemoEndo treatment group compared to background patient populations for different clinical variables as in panel C. Top axis in barplots corresponds to sample numbers.

B) Study layout

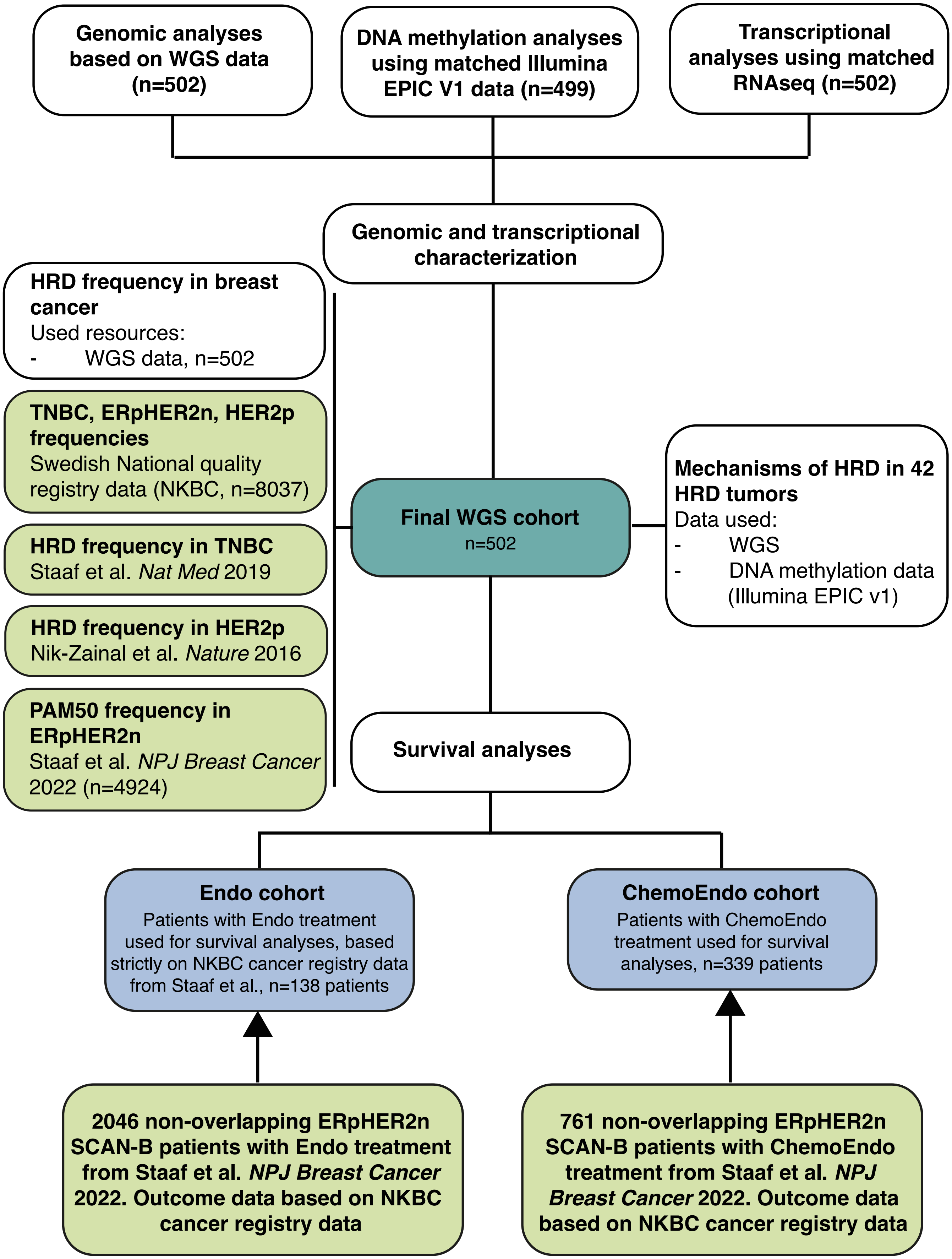

C)

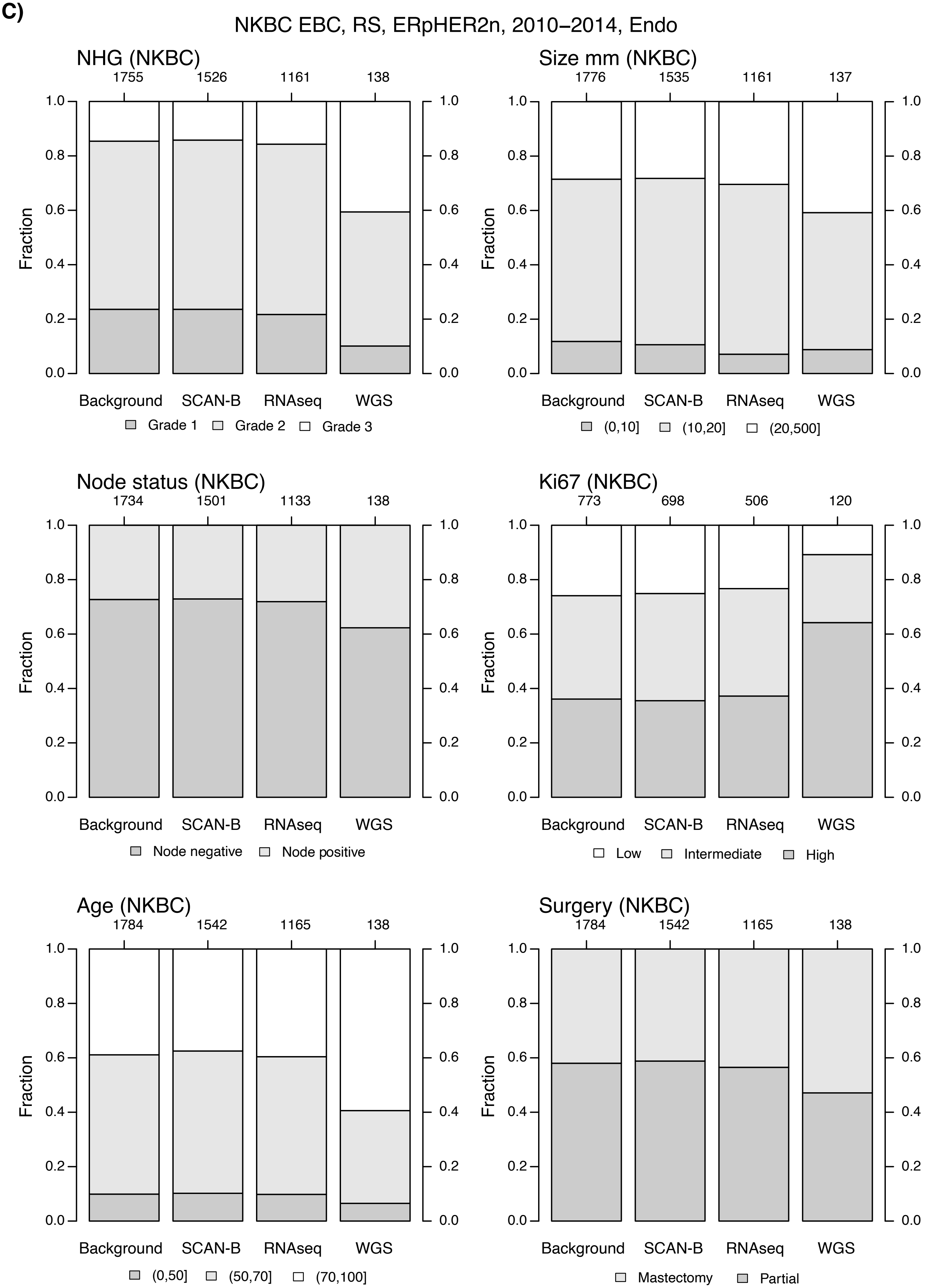

NKBC EBC, RS, ERpHER2n, 2010−2014, Endo
NHG (NKBC)
1755 1526 1161 138
Fraction
Background SCAN-B RNAseq WGS
Grade 1 Grade 2 Grade 3
Size mm (NKBC)
1776 1535 1161 137
Fraction
Background SCAN-B RNAseq WGS
(0,10] (10,20] (20,500]
Node status (NKBC)
1734 1501 1133 138
Fraction
Background SCAN-B RNAseq WGS
Node negative Node positive
Ki67 (NKBC)
773 698 506 120
Fraction
Background SCAN-B RNAseq WGS
Low Intermediate High
Age (NKBC)
1784 1542 1165 138
Fraction
Background SCAN-B RNAseq WGS
(0,50] (50,70] (70,100]
Surgery (NKBC)
1784 1542 1165 138
Fraction
Background SCAN-B RNAseq WGS
Mastectomy Partial

D)

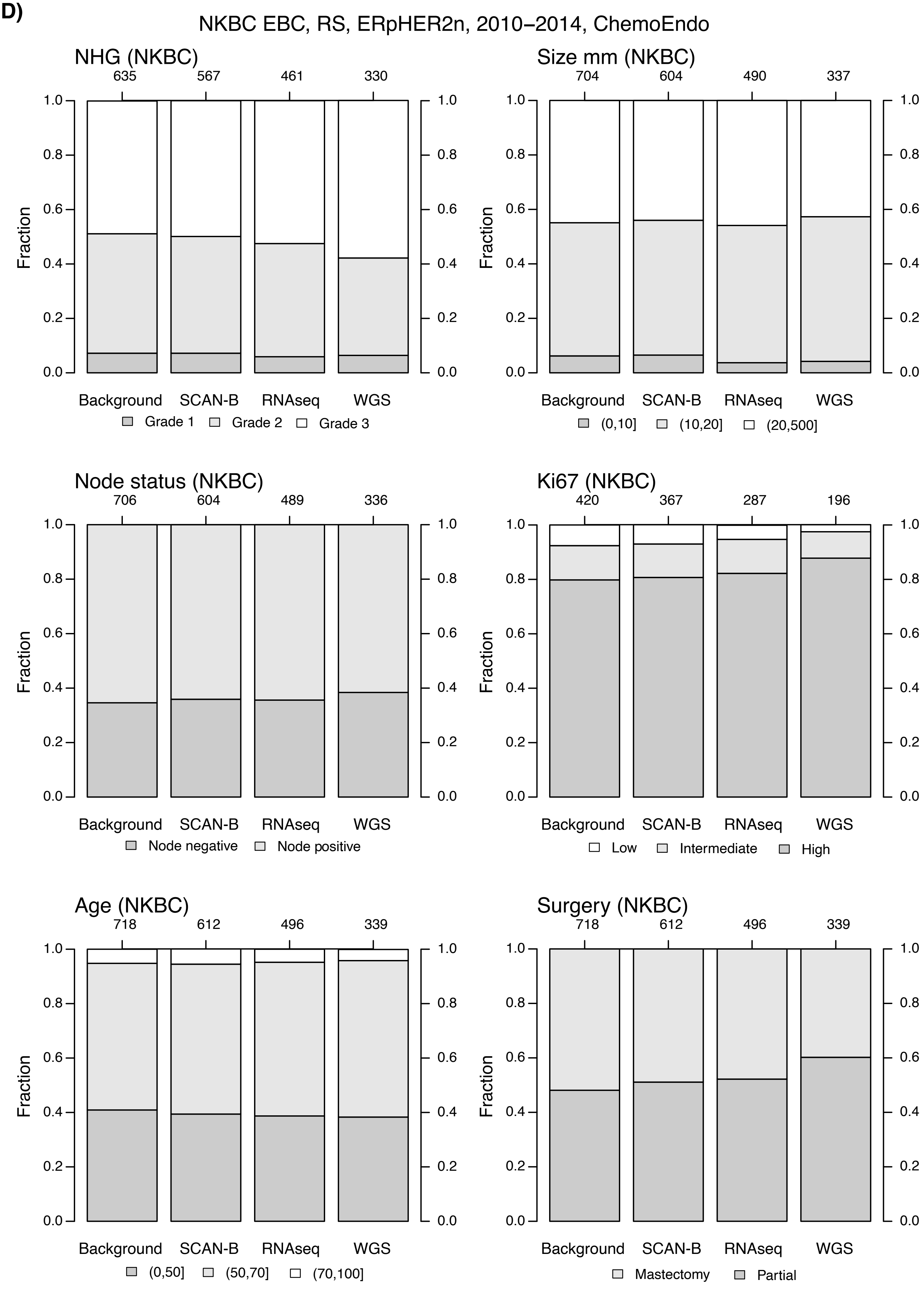

NKBC EBC, RS, ERpHER2n, 2010–2014, ChemoEndo
NHG (NKBC)
635 567 461 330
Fraction
Background SCAN-B RNAseq WGS
Grade 1 Grade 2 Grade 3
Size mm (NKBC)
704 604 490 337
(0,10] (10,20] (20,500]
Node status (NKBC)
706 604 489 336
Node negative Node positive
Ki67 (NKBC)
420 367 287 196
Low Intermediate High
Age (NKBC)
718 612 496 339
(0,50] (50,70] (70,100]
Surgery (NKBC)
718 612 496 339
Mastectomy Partial
