## Supplementary Figure S2 for "Homologous recombination deficiency in primary ER-positive and HER2-negative breast cancer"

A)

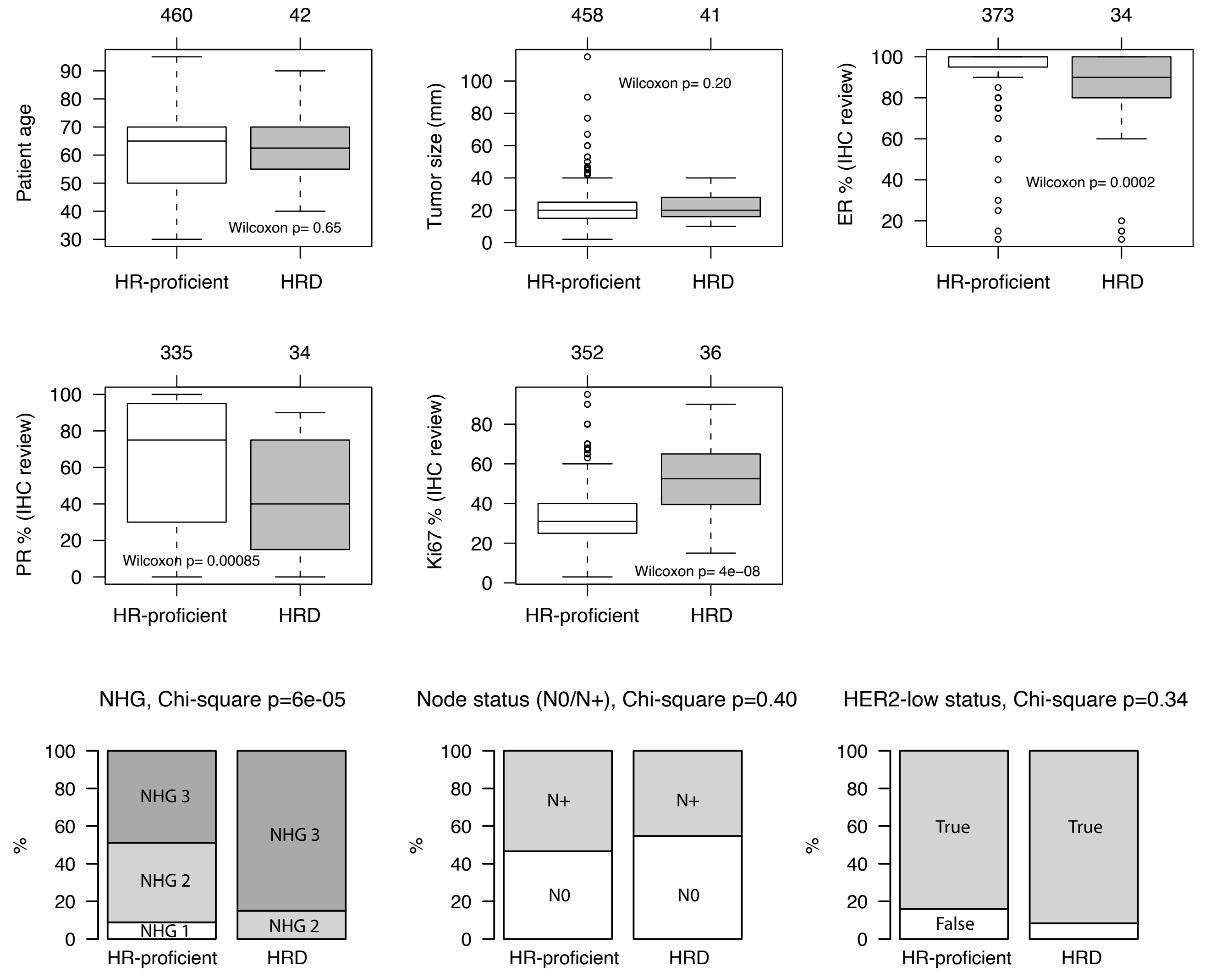


**Supplementary Figure S2. Clinicopathological characteristics of HRD and HR-proficient ERpHER2n tumors in 502 WGS analyzed patients divided by treatment status. (A)** Characteristics for all 502 patients. Not all patients have review data available. **(B)** Characteristics for 339 ChemoEndo treated patients. **(C)** Characteristics for 138 Endo treated patients. Not all patients have review data available. NHG: Nottingham grade index. N0: lymph node negative. N+: lymph node positive. Boxplot elements correspond to: i) center line = median, ii) box limits = upper and lower quartiles, iii) whiskers = 1.5x interquartile range. In boxplots, top axes indicate group sizes.

B)

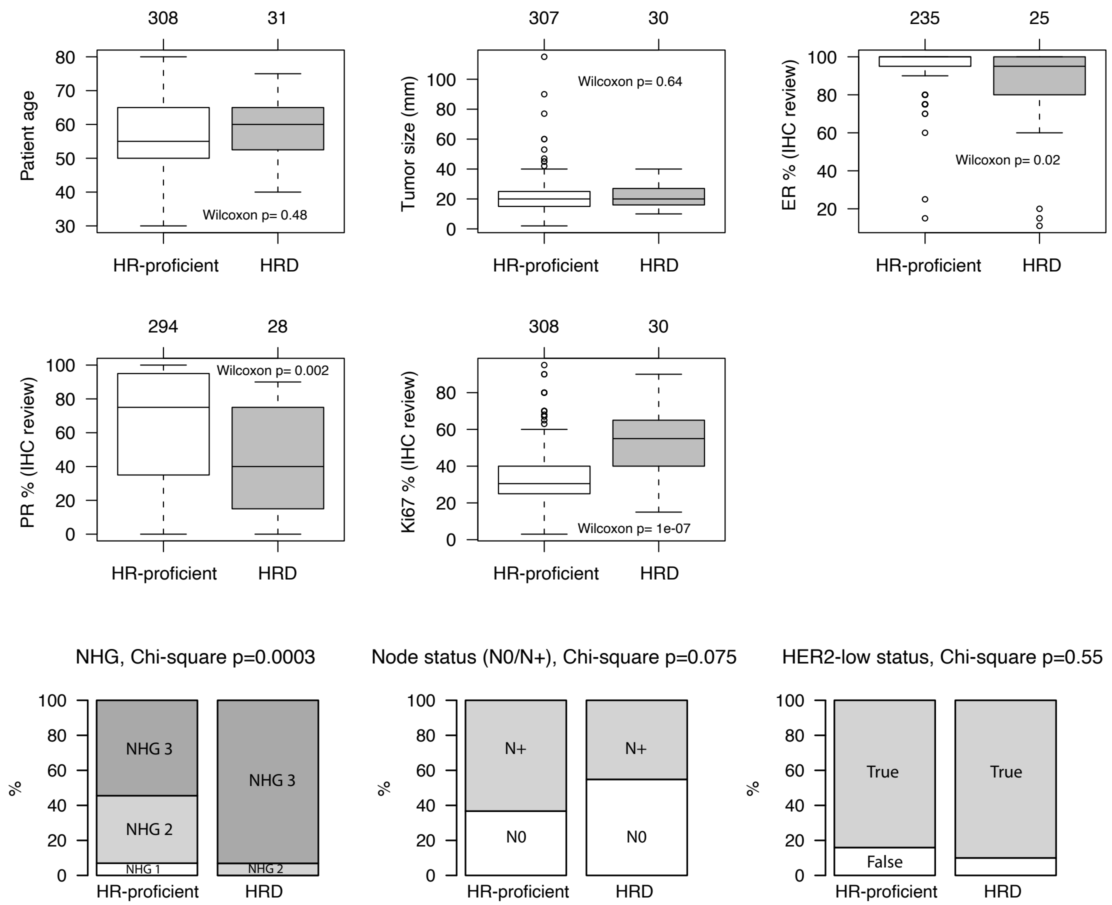
308
31
Patient age
Wilcoxon p= 0.48
HR-proficient
HRD
307
30
Tumor size (mm)
Wilcoxon p= 0.64
HR-proficient
HRD
235
25
ER % (IHC review)
Wilcoxon p= 0.02
HR-proficient
HRD
294
28
PR % (IHC review)
Wilcoxon p= 0.002
HR-proficient
HRD
308
30
Ki67 % (IHC review)
Wilcoxon p= 1e-07
HR-proficient
HRD
NHG, Chi-square p=0.0003
%
NHG 3
NHG 2
NHG 1
NHG 3
NHG 2
HR-proficient
HRD
Node status (N0/N+), Chi-square p=0.075
%
N+
N0
N+
N0
HR-proficient
HRD
HER2-low status, Chi-square p=0.55
%
True
False
True
HR-proficient
HRD

C)

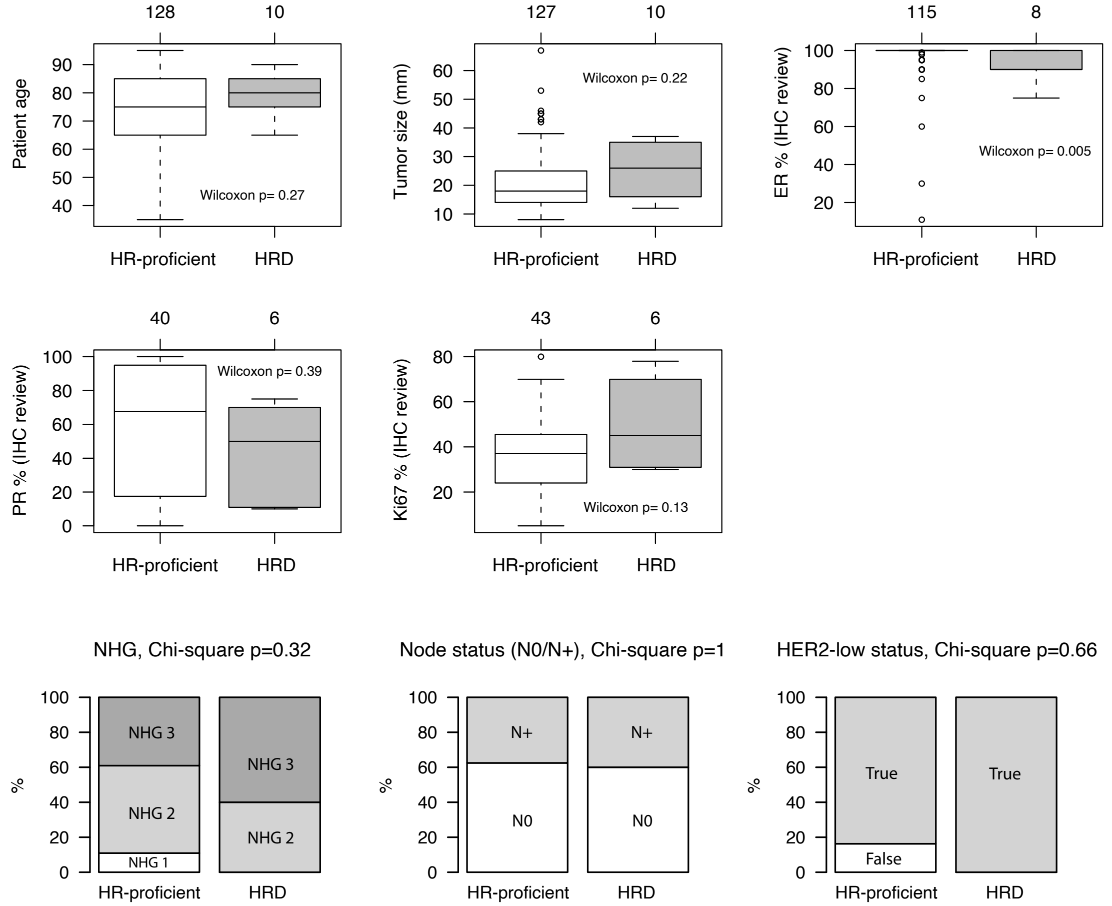
128
10
Patient age
Wilcoxon p= 0.27
HR-proficient
HRD
127
10
Tumor size (mm)
Wilcoxon p= 0.22
HR-proficient
HRD
115
8
ER % (IHC review)
Wilcoxon p= 0.005
HR-proficient
HRD
40
6
PR % (IHC review)
Wilcoxon p= 0.39
HR-proficient
HRD
43
6
Ki67 % (IHC review)
Wilcoxon p= 0.13
HR-proficient
HRD
NHG, Chi-square p=0.32
%
NHG 3
NHG 2
NHG 1
NHG 3
NHG 2
HR-proficient
HRD
Node status (N0/N+), Chi-square p=1
%
N+
N0
N+
N0
HR-proficient
HRD
HER2-low status, Chi-square p=0.66
%
True
False
True
HR-proficient
HRD
