## Supplementary Figure S3 for "Homologous recombination deficiency in primary ER-positive and HER2-negative breast cancer"

### A) PAM50 Basal

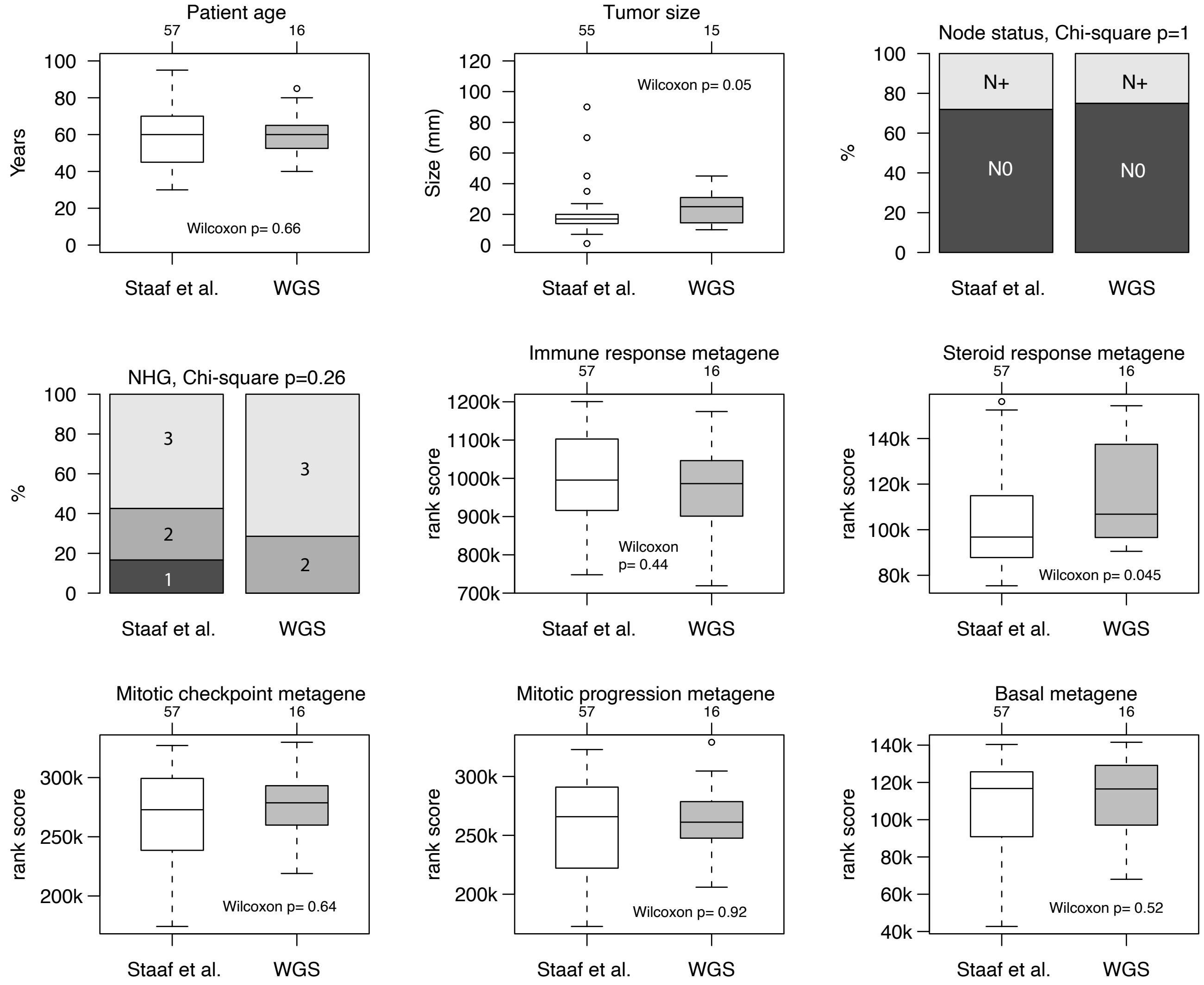


**Supplementary Figure S3. Representativity of PAM50 subtypes in the ERpHER2n WGS cohort versus PAM50 subtypes in 4427 unrelated ERpHER2n tumors from the study by Staaf et al. (NPJ Breast Cancer 2022) regarding clinicopathological characteristics and expression of biological mRNA metagenes from Fredlund et al. (BCR 2012).** The Chi-square test was used to test statistical significance for categorical data, whereas Wilcoxon's test was used for continuous data and rank-scores. (**A**) PAM50 Basal. (**B**) PAM50 HER2E. (**C**) PAM50 LumA. (**D**) PAM50 LumB. (**E**) PAM50 Normal. While certain characteristics in panel D (LumB) are significant it should be noted that the larger group sizes here can cause smaller differences to become statistically significant (e.g. age and nodal status). NHG: Nottingham grade index (grade 1, 2, 3). N0: lymph node negative. N+: lymph node positive.

### B) PAM50 HER2E

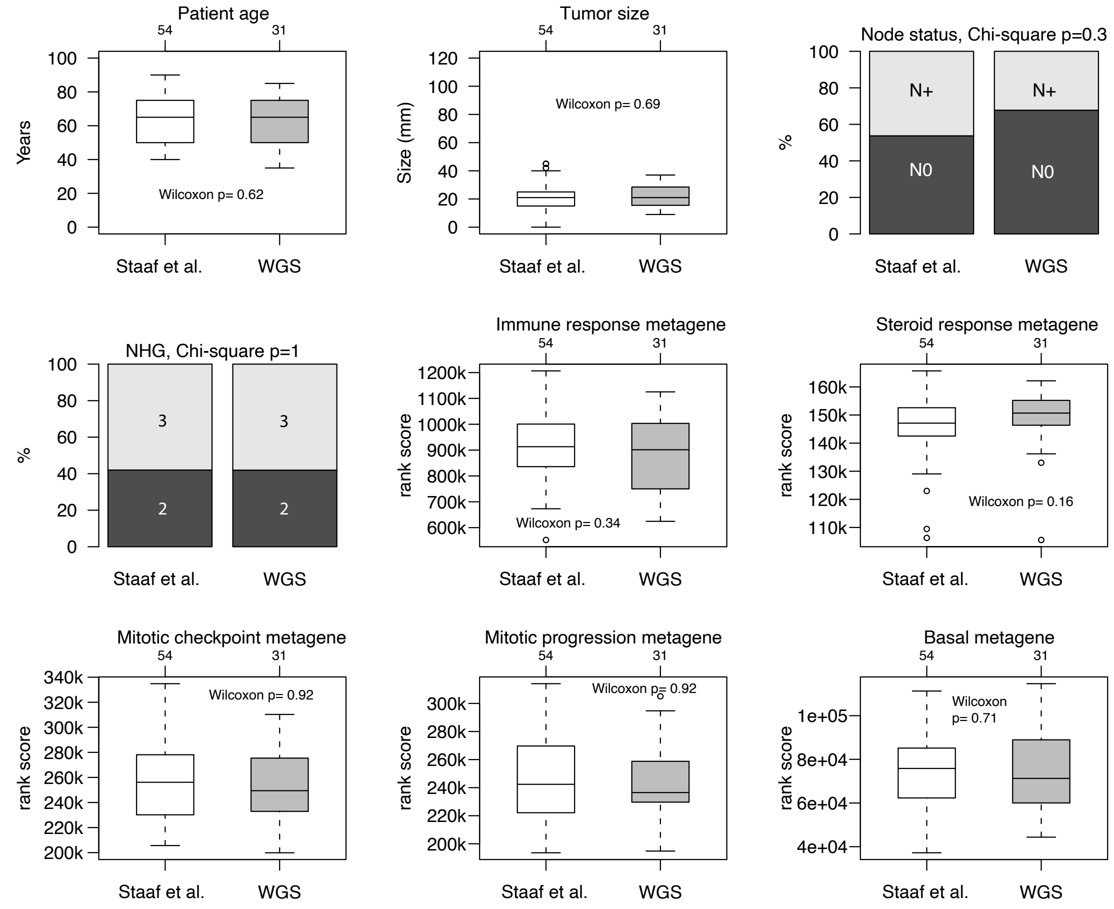


### C) PAM50 LumA

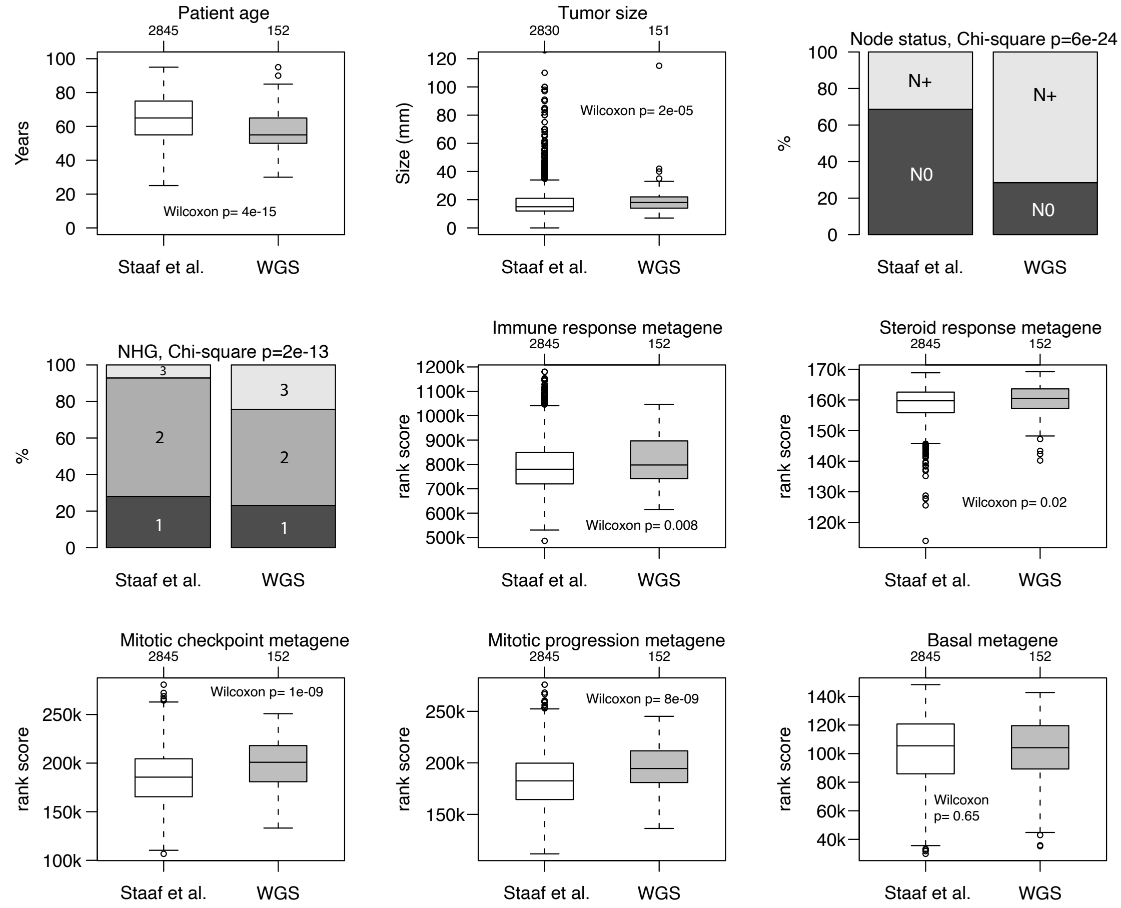

### D) PAM50 LumB

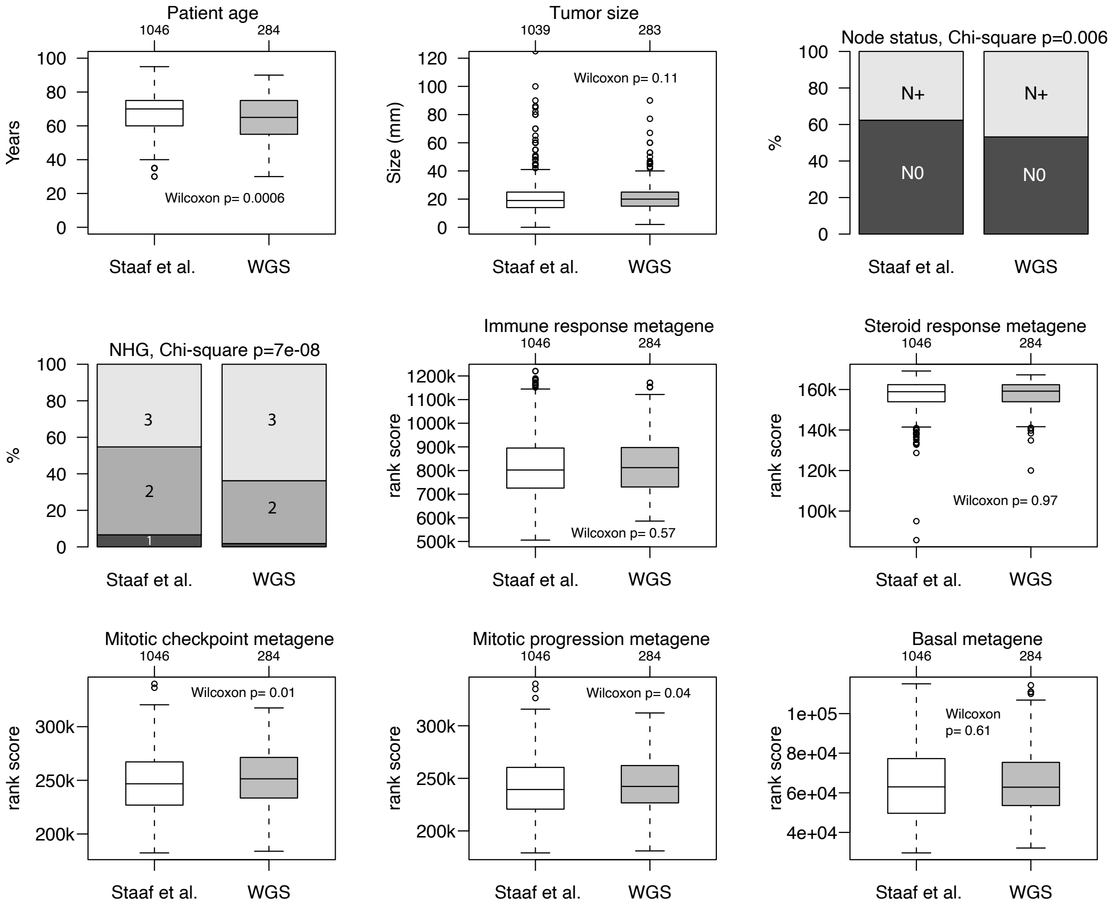


### E) PAM50 Normal

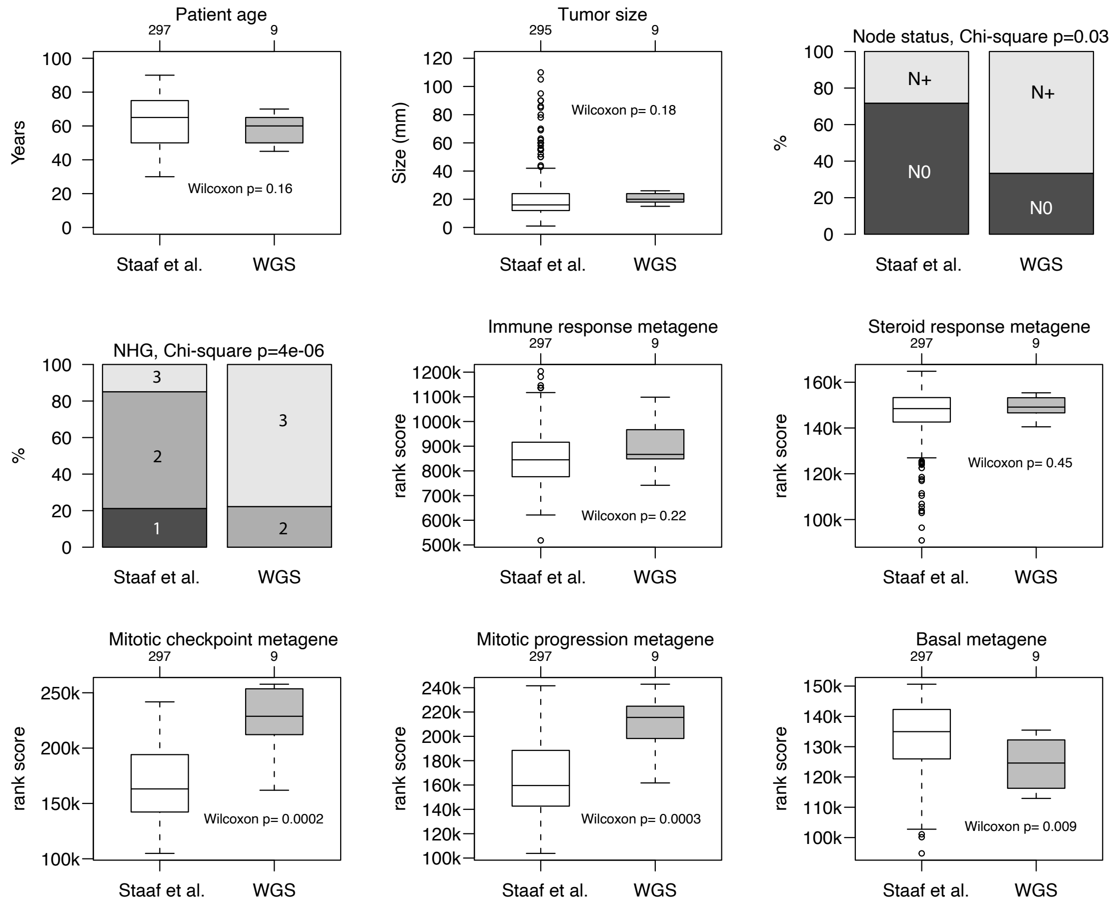
