## Supplementary Figure S4 for "Homologous recombination deficiency in primary ER-positive and HER2-negative breast cancer"

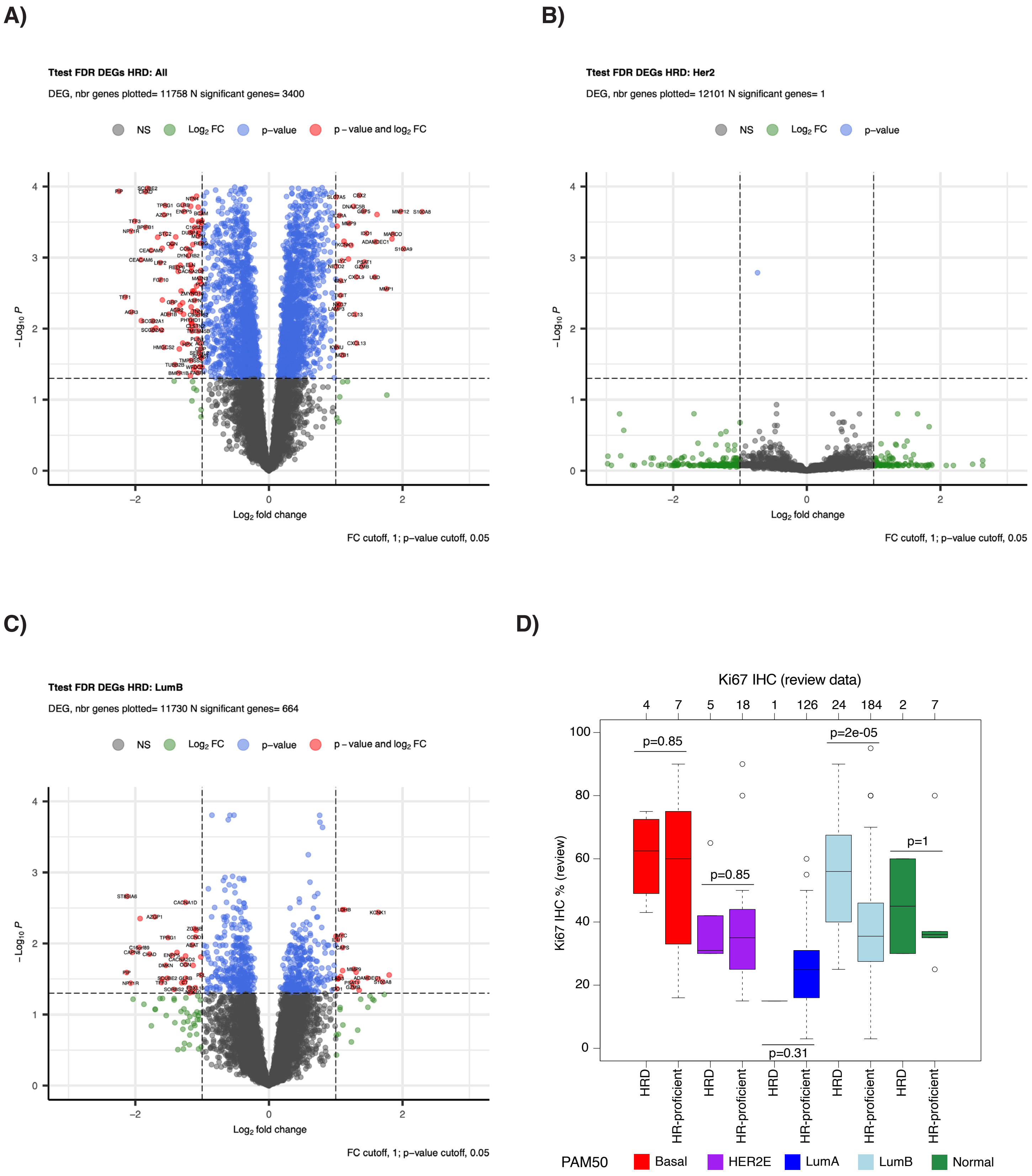


**Supplementary Figure S4. Gene expression analysis of ERpHER2n tumors with respect to HRD status.** (**A**) Volcano plot showing the results of the differential gene expression analysis for HRD-status in all 502 tumors. (**B**) Volcano plot showing the results of the differential gene expression analysis for HRD-status in HER2E tumors. (**C**) Volcano plot showing the results of the differential gene expression analysis for HRD-status in LumB tumors. In A-C, the fold change (FC) cut-off was 1, and the p-value FDR cut-off 0.05. (**D**) Ki67 IHC expression based on clinical review data for tumors stratified by PAM50 and HRD status, showing higher Ki67 levels in HRD LumB tumors. Two-sided p-values calculated using Wilcoxon's test.
