## Supplementary Figure S5 for "Homologous recombination deficiency in primary ER-positive and HER2-negative breast cancer"

**A)**

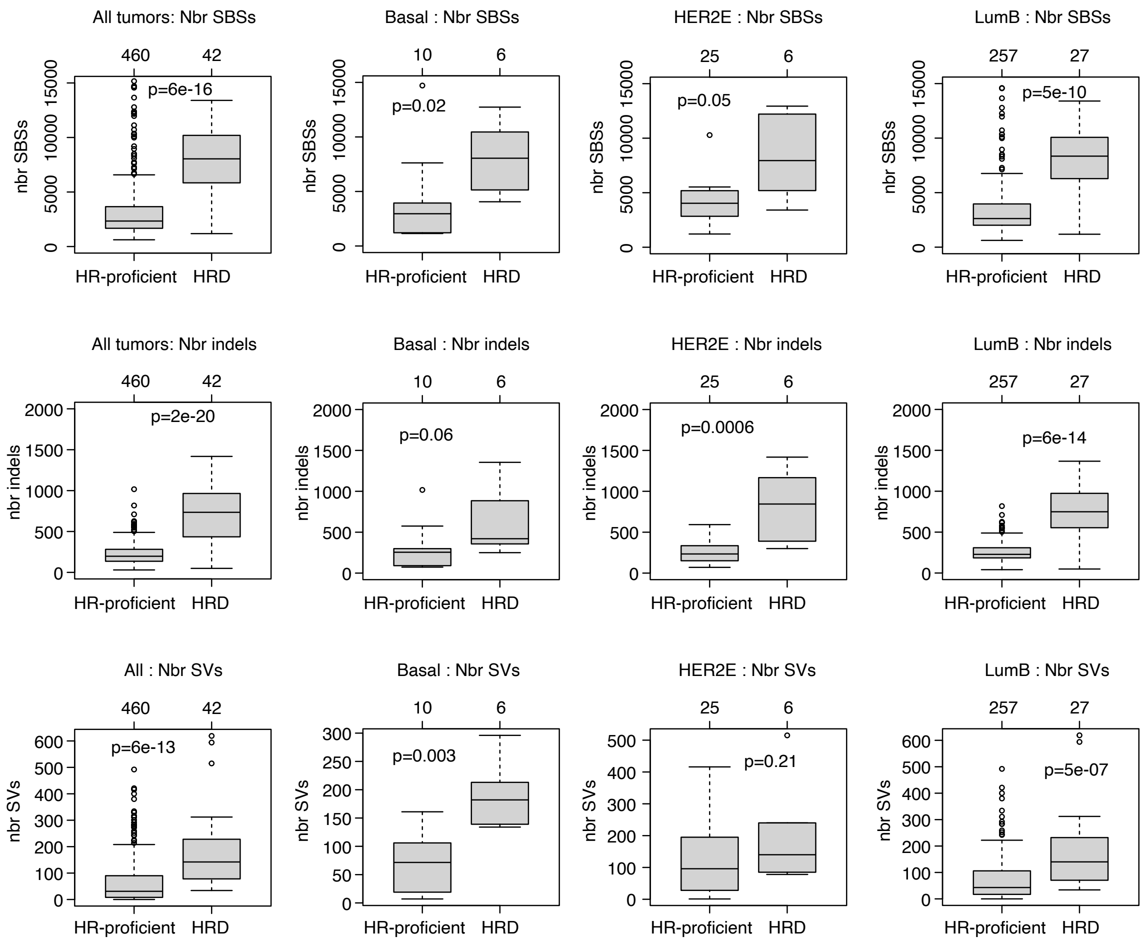


**Supplementary Figure S5. Genetic features of HRD ERpHER2n tumors. (A)** Number of single base substitutions (SBSs), indels, and structural rearrangements (SVs) in HRD versus HR-proficient tumors stratified by PAM50 subtype (LumA and Normal excluded due to few HRD cases). Two-sided p-values are calculated using Wilcoxon's test. **(B)** Exposure to SBS signatures for all tumors, Basal, HER2E, and LumB. Only SBS1, SBS3, SBS5, SBS8, and SBS2/13 are shown as these were the signatures with significant differences between groups. Two-sided p-values are calculated using Wilcoxon's test. **(C)** Exposure to SV signatures for all tumors, Basal, HER2E, and LumB subtypes. Only signatures significant or borderline nonsignificant are shown. Two-sided p-values are calculated using Wilcoxon's test. **(D)** Exposure to indel signatures for all tumors, Basal, HER2E, and LumB subtypes. Only signatures with significant differences are shown. Two-sided p-values are calculated using Wilcoxon's test. **(E)** WGS estimated tumor ploidy, fraction of the genome altered by copy number alterations (CN_FGA), fraction of the genome altered by LOH (LOH_FGA), and number of ASCAT segments on chromosome 1-22 per tumor called as gain or loss for all tumors, Basal, HER2E, and LumB subtypes. **(F)** Barplot of FDR adjusted T-test values (-log10 transformed) for 25 copy number signatures (CN1-25) between HR-proficient and HRD tumors stratified by PAM50 subtype. E.g. for CN1, significant p-values (FDR>0.05) was found for the comparison of HRD vs HR-proficient tumors in the total cohort and in LumB tumors specifically. **(G)** Left: frequency of gene driver events based on somatic SBSs and indels in PAM50 Basal tumors stratified by HRD status. Right: frequency of gene driver events based on somatic structural rearrangements (SVs) in PAM50 Basal tumors stratified by HRD status. Only genes detected in at least two tumors in at least one group are shown. **(H)** As in G but for PAM50 HER2E tumors. (**I**) Principal component analysis (PCA) of tumor purity adjusted DNA methylation data (beta values) for three different CpG contexts in all tumors and PAM50 subtypes (LumA excluded as only one tumor was HRD). For each context, the 5000 most variant CpGs in each subgroup of samples were selected for PCA. Black dots represent tumors with an HRDetect HRD classification. First two principal components (PC1 and PC2) shown. (**J**) PCA of tumor purity adjusted DNA methylation data (beta values) for three different CpG contexts in LumB HRD tumors. For each context, the 5000 most variant CpGs in the subgroup were selected for PCA. Samples are colored based on their proposed HRD inactivation mechanism. First two principal components (PC1 and PC2) shown.

B)

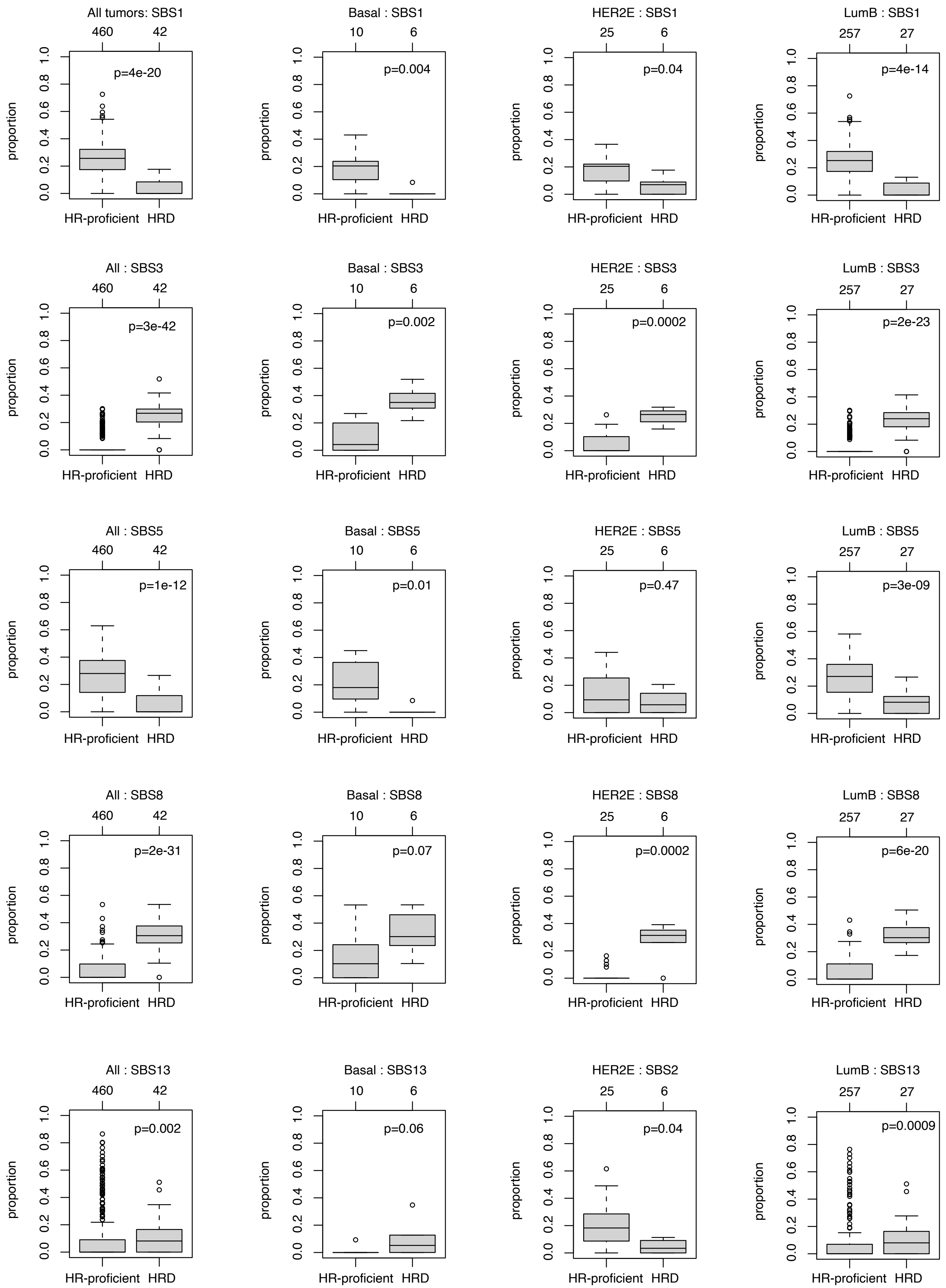
All tumors: SBS1
460 42
p=4e-20
Basal : SBS1
10 6
p=0.004
HER2E : SBS1
25 6
p=0.04
LumB : SBS1
257 27
p=4e-14
All : SBS3
460 42
p=3e-42
Basal : SBS3
10 6
p=0.002
HER2E : SBS3
25 6
p=0.0002
LumB : SBS3
257 27
p=2e-23
All : SBS5
460 42
p=1e-12
Basal : SBS5
10 6
p=0.01
HER2E : SBS5
25 6
p=0.47
LumB : SBS5
257 27
p=3e-09
All : SBS8
460 42
p=2e-31
Basal : SBS8
10 6
p=0.07
HER2E : SBS8
25 6
p=0.0002
LumB : SBS8
257 27
p=6e-20
All : SBS13
460 42
p=0.002
Basal : SBS13
10 6
p=0.06
HER2E : SBS2
25 6
p=0.04
LumB : SBS13
257 27
p=0.0009
proportion
0.0 0.2 0.4 0.6 0.8 1.0
HR-proficient
HRD

C)

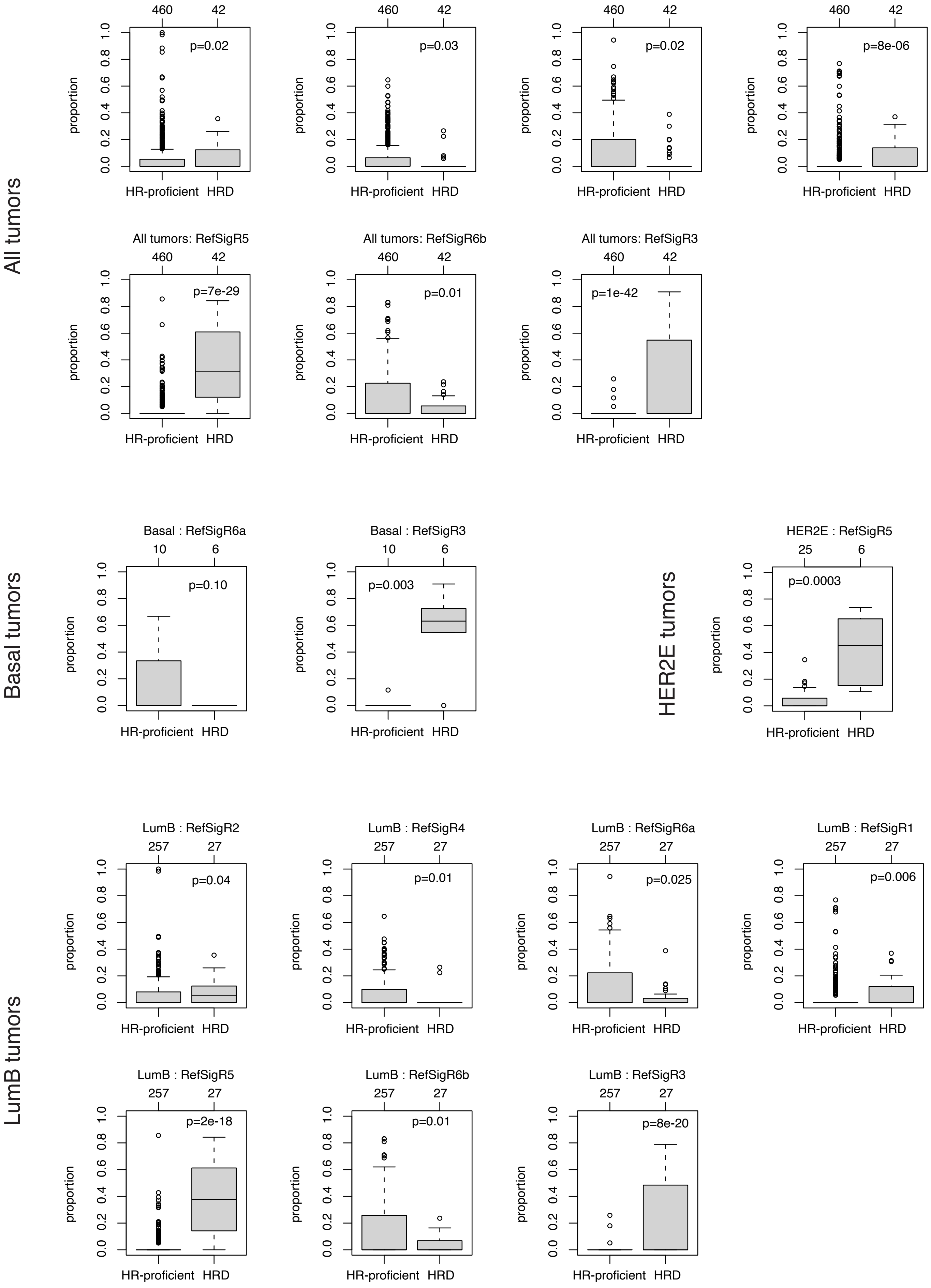
All tumors
All tumors: RefSigR2
p=0.02
All tumors: RefSigR4
p=0.03
All tumors: RefSigR6a
p=0.02
All tumors: RefSigR1
p=8e-06
All tumors: RefSigR5
p=7e-29
All tumors: RefSigR6b
p=0.01
All tumors: RefSigR3
p=1e-42
460
42
proportion
HR-proficient
HRD
Basal tumors
Basal : RefSigR6a
p=0.10
Basal : RefSigR3
p=0.003
10
6
HER2E tumors
HER2E : RefSigR5
p=0.0003
25
6
LumB tumors
LumB : RefSigR2
p=0.04
LumB : RefSigR4
p=0.01
LumB : RefSigR6a
p=0.025
LumB : RefSigR1
p=0.006
LumB : RefSigR5
p=2e-18
LumB : RefSigR6b
p=0.01
LumB : RefSigR3
p=8e-20
257
27

D)

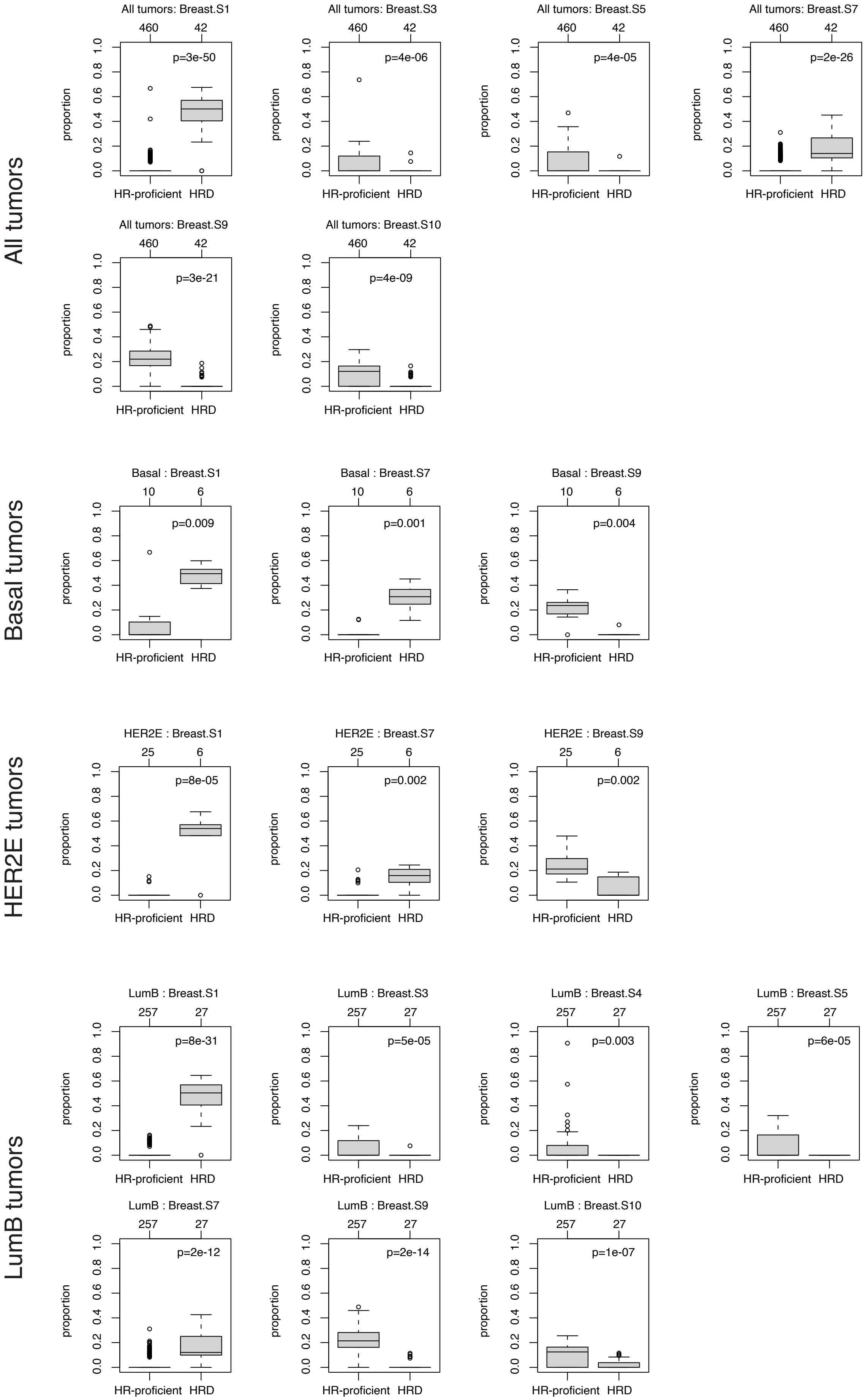
All tumors
All tumors: Breast.S1
460 42
p=3e-50
All tumors: Breast.S3
460 42
p=4e-06
All tumors: Breast.S5
460 42
p=4e-05
All tumors: Breast.S7
460 42
p=2e-26
All tumors: Breast.S9
460 42
p=3e-21
All tumors: Breast.S10
460 42
p=4e-09
Basal tumors
Basal : Breast.S1
10 6
p=0.009
Basal : Breast.S7
10 6
p=0.001
Basal : Breast.S9
10 6
p=0.004
HER2E tumors
HER2E : Breast.S1
25 6
p=8e-05
HER2E : Breast.S7
25 6
p=0.002
HER2E : Breast.S9
25 6
p=0.002
LumB tumors
LumB : Breast.S1
257 27
p=8e-31
LumB : Breast.S3
257 27
p=5e-05
LumB : Breast.S4
257 27
p=0.003
LumB : Breast.S5
257 27
p=6e-05
LumB : Breast.S7
257 27
p=2e-12
LumB : Breast.S9
257 27
p=2e-14
LumB : Breast.S10
257 27
p=1e-07
proportion
0.0 0.2 0.4 0.6 0.8 1.0
HR-proficient HRD

**E)**

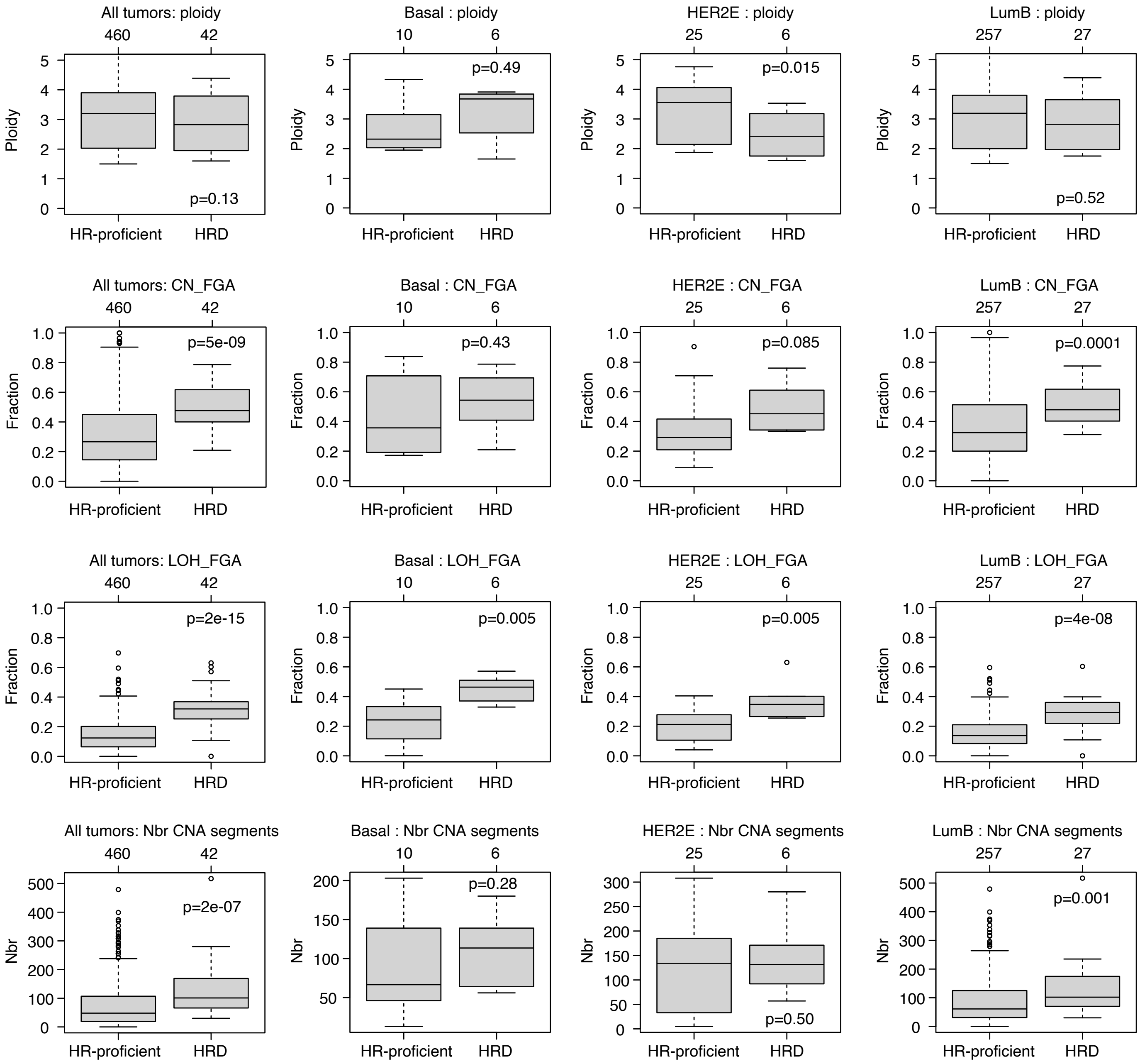
All tumors: ploidy
460
42
Ploidy
p=0.13
HR-proficient
HRD
Basal : ploidy
10
6
p=0.49
HER2E : ploidy
25
6
p=0.015
LumB : ploidy
257
27
p=0.52
All tumors: CN_FGA
Fraction
p=5e-09
Basal : CN_FGA
p=0.43
HER2E : CN_FGA
p=0.085
LumB : CN_FGA
p=0.0001
All tumors: LOH_FGA
p=2e-15
Basal : LOH_FGA
p=0.005
HER2E : LOH_FGA
p=0.005
LumB : LOH_FGA
p=4e-08
All tumors: Nbr CNA segments
Nbr
p=2e-07
Basal : Nbr CNA segments
p=0.28
HER2E : Nbr CNA segments
p=0.50
LumB : Nbr CNA segments
p=0.001

**F)**

CN signature proportions vs HRDetect

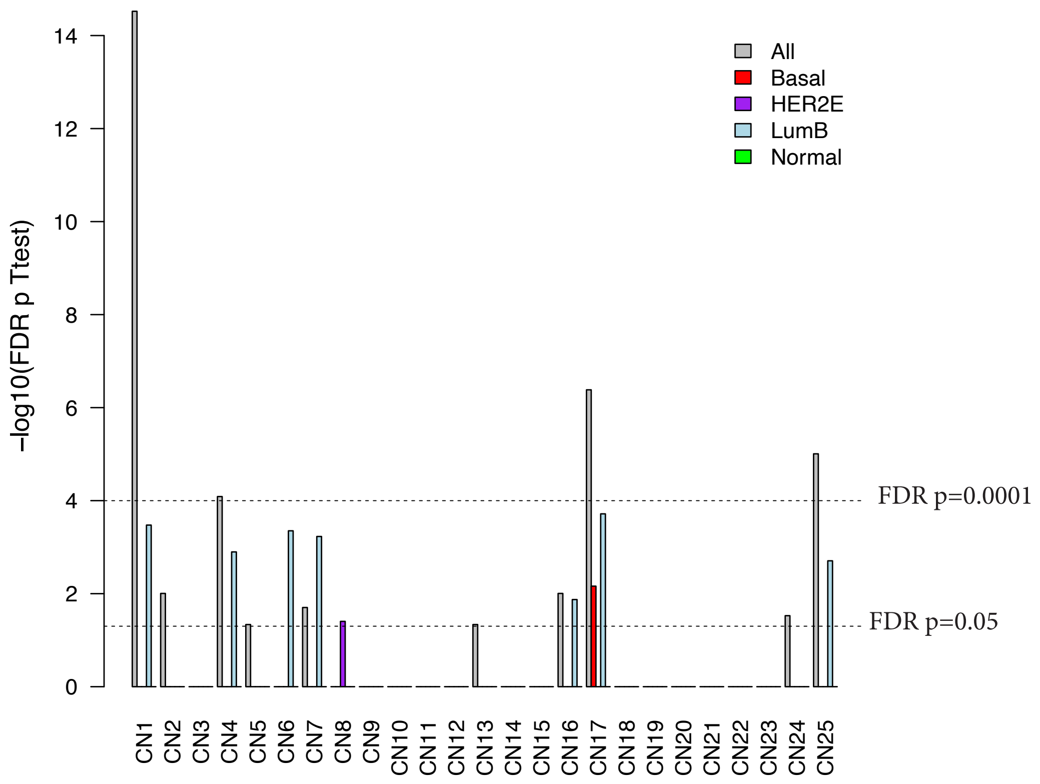


**G)** PAM50 Basal

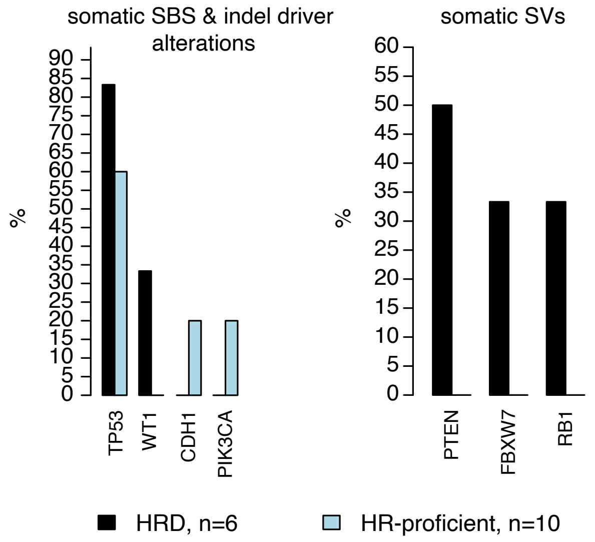


**H)** PAM50 HER2E

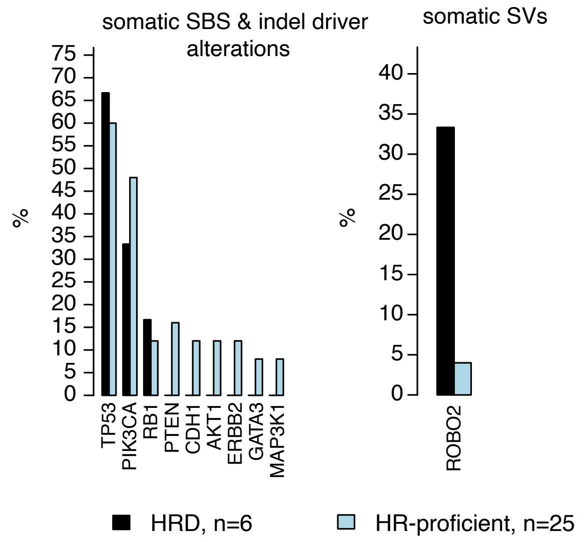

**I)**

### distal-ATAC

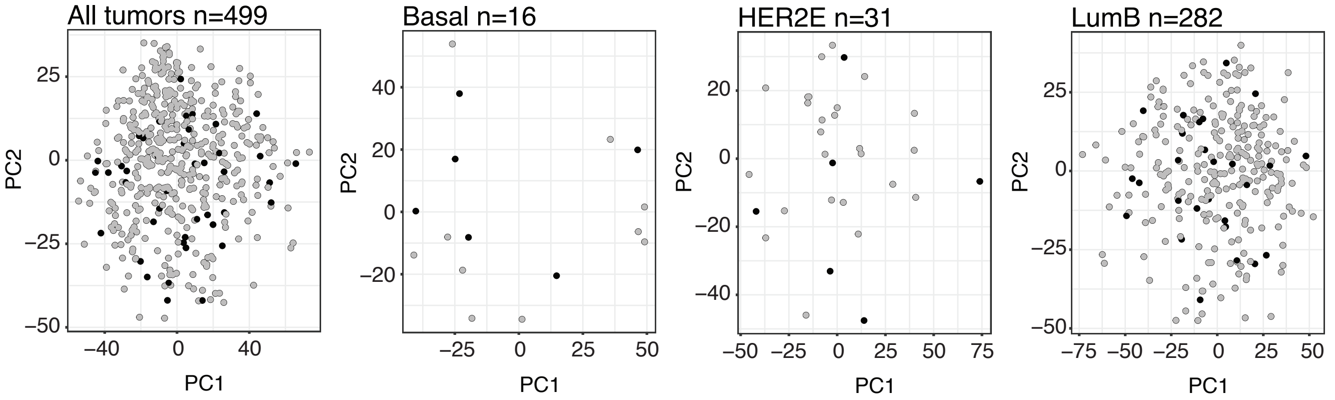


### proximal-ATAC

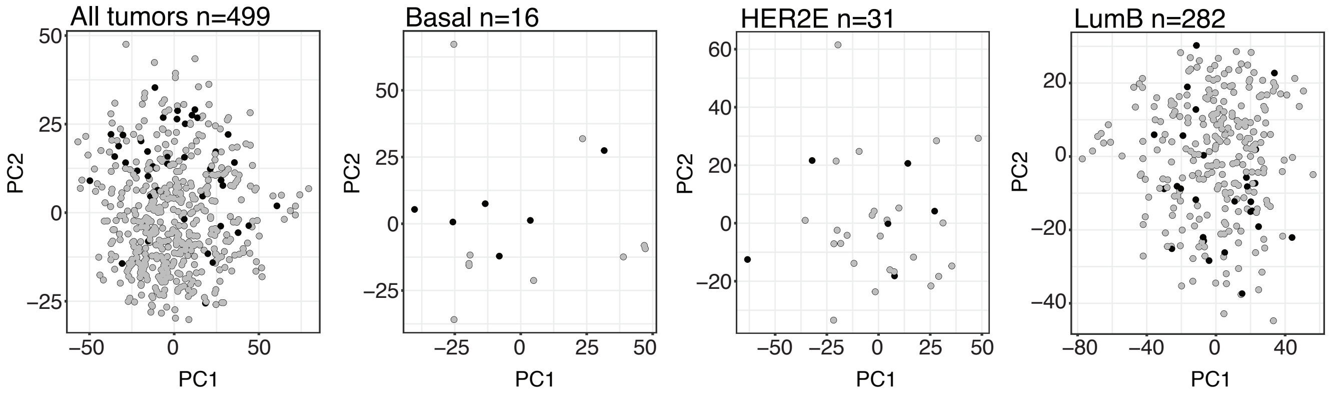


### promoter-ATAC

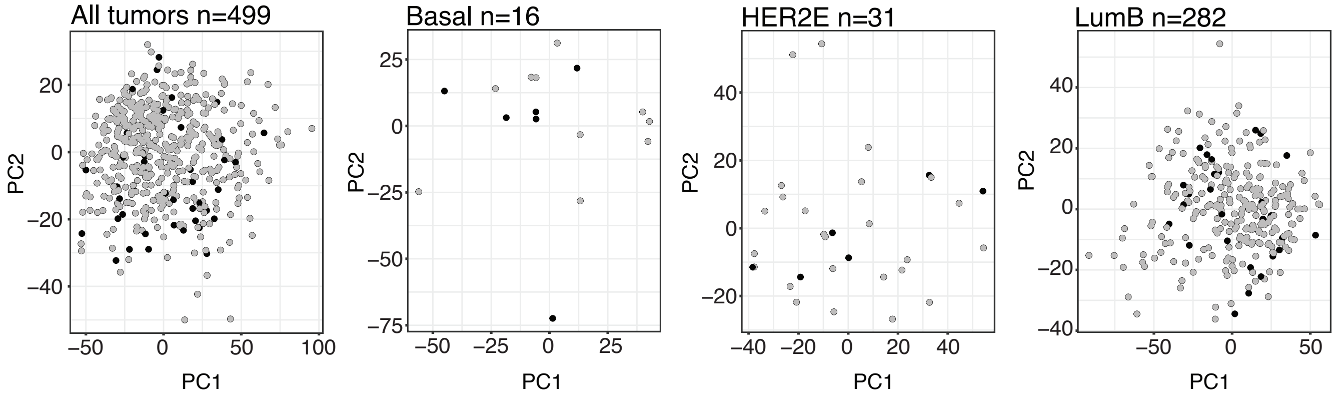

J)

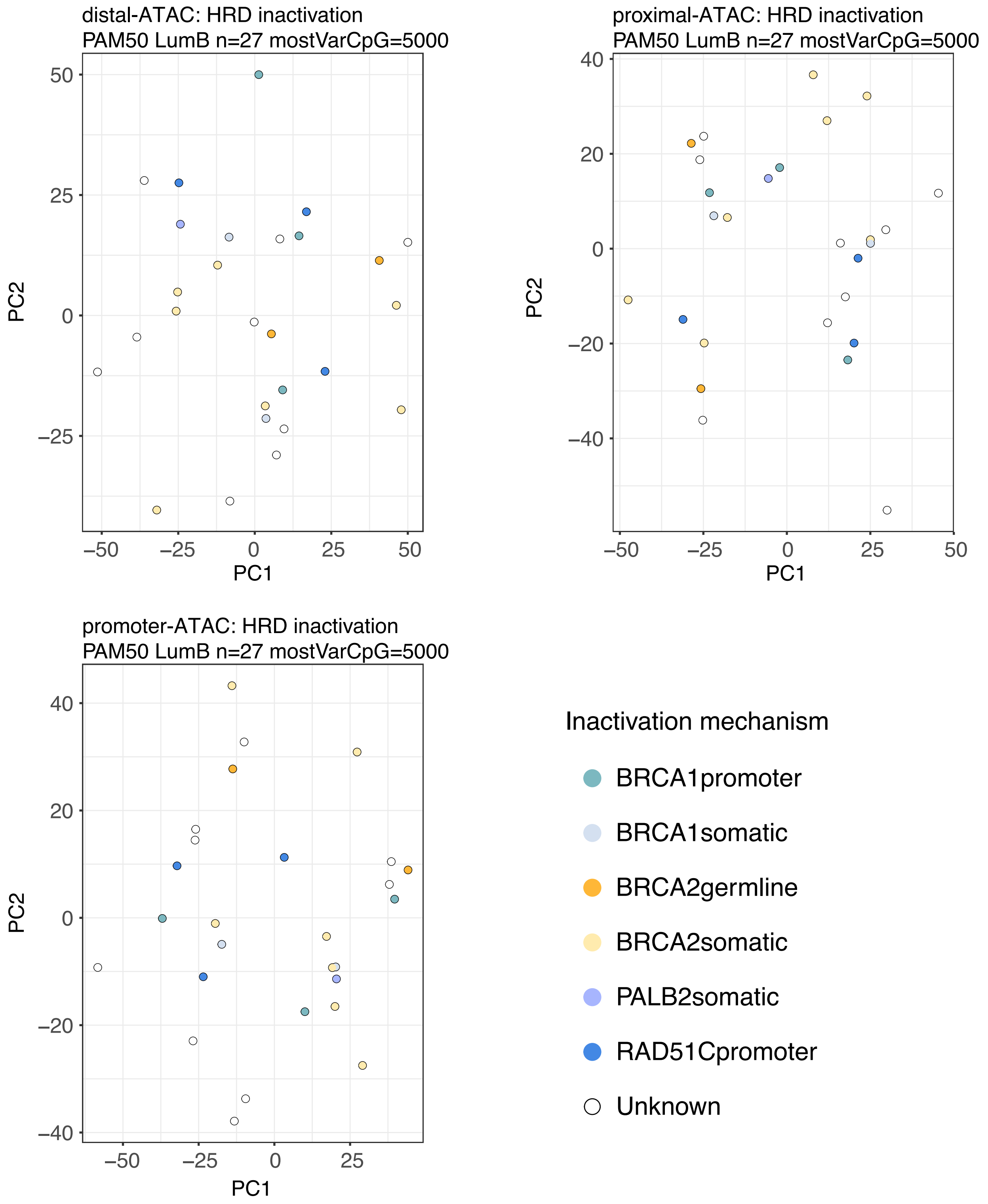
distal-ATAC: HRD inactivation
PAM50 LumB n=27 mostVarCpG=5000
PC2
PC1
50
25
0
−25
−50
−25
0
25
50
proximal-ATAC: HRD inactivation
PAM50 LumB n=27 mostVarCpG=5000
PC2
PC1
40
20
0
−20
−40
−50
−25
0
25
50
promoter-ATAC: HRD inactivation
PAM50 LumB n=27 mostVarCpG=5000
PC2
PC1
40
20
0
−20
−40
−50
−25
0
25
Inactivation mechanism
BRCA1promoter
BRCA1somatic
BRCA2germline
BRCA2somatic
PALB2somatic
RAD51Cpromoter
Unknown
