## Supplementary Figure S6 for "Homologous recombination deficiency in primary ER-positive and HER2-negative breast cancer"

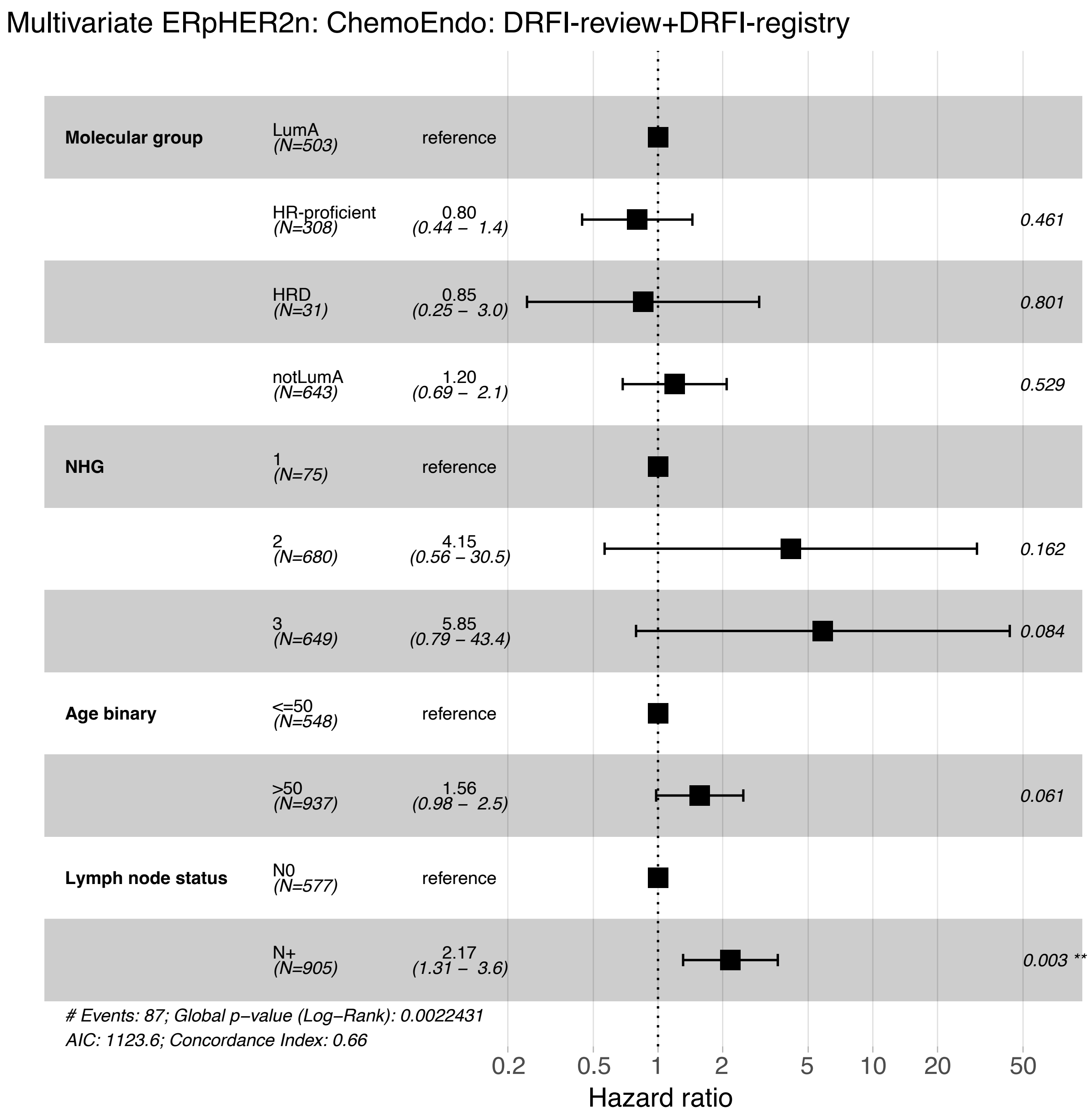


**Supplementary Figure S6. Multivariate Cox regression in ChemoEndo ERpHER2n BC.** Forest plot of hazard ratios and 95% confidence intervals for a multivariate Cox regression model including a merged molecular class (PAM50 LumA yes/no and HRD status), Nottingham grade index (NHG), binary age, and binary lymph node status in ChemoEndo treated ERpHER2n patients using distant relapse-free interval (DRFI) as clinical endpoint. In this analysis, WGS-analyzed ChemoEndo treated patients was merged with 761 non-overlapping ChemoEndo treated patients with unknown HRD status stratified by their PAM50 LumA status (LumA or not-LumA). For the 761 patients the DRFI data based on cancer registry data from, while for the WGS analyzed samples clinical review DRFI data was used as endpoint.
